# UCF-MultiOrgan-Path:A Benchmark Dataset of Histopathologic Images for Deep Learning-Based Organ Classification

**DOI:** 10.1101/2024.11.05.24316736

**Authors:** Md Sanzid Bin Hossain, Yelena Piazza, Jacob Braun, Anthony Bilic, Michael Hsieh, Samir Fouissi, Alexander Borowsky, Hatem Kaseb, Chaithanya Renduchintala, Amoy Fraser, Britney-Ann Wray, Chen Chen, Liqiang Wang, Mujtaba Husain, Dexter Hadley

## Abstract

A pathologist typically diagnoses tissue samples by examining glass slides under a light microscope. The entire tissue specimen can be stored digitally as a Whole Slide Image (WSI) for further analysis. However, managing and diagnosing large numbers of images manually is time-consuming and requires specialized expertise. Consequently, computer-aided diagnosis of these pathology images is an active research area, with deep learning showing promise in disease classification and cancer cell segmentation. Robust deep learning models need many annotated images, but public datasets are limited, often constrained to specific organs, cancer types, or binary classifications, which limits generalizability. To address this, we introduce the UCF multi-organ histopathologic (UCF-MultiOrgan-Path) dataset, containing 977 WSIs from cadaver tissues across 15 organ classes, including lung, kidney, liver, and pancreas. This dataset includes ∼2.38 million patches of 512×512 pixels. For technical validation, we provide patch-based and slide-based approaches for patch- and slide-level classification. Our dataset, containing millions of patches, can serve as a benchmark for training and validating deep learning models in multi-organ classification.

## Background & Summary

Histopathology, the study of tissues at the microscopic level, is an important component in disease diagnosis and cancer detection^1–5^. Traditionally, a pathologist examines stained tissue specimens under a microscope to identify abnormalities and make diagnoses^6^. However, with the emergence of digital pathology^7^, whole slide image (WSI) has become increasingly popular, allowing for the digitization and storage of entire tissue specimens for further analysis^8^. This transformation has greatly expanded histopathology’s practical utility, including enhanced teaching efficiency, reduced diagnosis costs^9^, and improved research capabilities^10,11^.

Despite these advances, manually diagnosing and analyzing hundreds of WSIs is a labor-intensive and challenging task^10^, requiring a high level of digital pathology expertise and thorough file management. To alleviate these challenges, significant research has focused on developing computer-aided diagnosis (CAD) systems for digitally acquired pathology images^12,13^. Among these advancements, deep learning, a subfield of machine learning, has emerged as a powerful tool due to its ability to learn complex patterns and features from large datasets^14,15^. A major challenge in developing deep learning models for histopathologic image analysis is the scarcity of large, annotated public datasets that accurately represent real-world clinical scenarios^16–19^. While existing datasets are often curated for specific machine learning applications, they frequently lack the diversity and complexity of real clinical data. The performance of deep learning models heavily depends on the availability of such diverse and accurately labeled datasets^20^. The FAIR principles (Findability, Accessibility, Interoperability, and Reusability) were developed to address this gap and foster collaboration within the scientific community^21,22^. However, many existing datasets remain limited in size, diversity, and annotations, hindering the development of robust and generalizable models^16–20,23,24^. To address these limitations, we have curated a public dataset named UCF-MultiOrgan-Path that provides a more realistic representation of clinical data. This dataset includes a large number of histopathologic WSIs collected from cadavers during medical school education at the University of Central Florida (UCF) over the course of 10 years (2010-2019). By spanning a decade, it captures a wide variety of patient cases, different types of diseases, and variations in causes of death, reflecting the natural variability and changes in medical practices over time. Our dataset contains WSIs from 15 organs, such as the lung, kidney, liver, pancreas, and others, reflecting the variety and complexity of cases encountered in real clinical settings. It offers important knowledge on histological structures and additionally can act as an outstanding educational resource, further showcasing its high quality and usefulness.

Table 1 compares existing histopathologic datasets, including their statistics and intended purposes, with our UCF-MultiOrgan-Path dataset. The UCF-MultiOrgan-Path dataset offers significant advantages over other public datasets in terms of dataset size, number of classes, and purpose. With 977 WSIs and approximately 2.38 million patches representing 15 organ classes, UCF-MultiOrgan-Path provides a broader scope for deep learning classification benchmarks in the context of multi-class pathology analysis. In contrast, other datasets such as Kimia Path24^25^ and Kimia Path24C^26^ are highly specialized, focusing on the classification of texture patterns from just 24 WSIs, restricting their applicability to broader tasks. Similarly, the Atlas of Digital Pathology^27^ includes 100 WSIs with a focus on multi-label classification across 57 categories but with only 17,688 patches, which is notably smaller compared to the extensive number of patches provided by UCF-MultiOrgan-Path. Datasets such as CAMELYON16/17^28–30^ and PatchCamelyon^33^ are constrained by their binary classification tasks (e.g., metastasis vs. no metastasis in lymph nodes), which reduces their utility as benchmarks for evaluating complex, multi-class scenarios in deep learning models. Similarly, TCGA^31^ offers a substantial number of tissue slides across various cancers, yet its narrow focus limits its effectiveness as a benchmark for evaluating models on a more diverse set of histopathological classes. Meanwhile, BACH^32^ focuses specifically on breast cancer classification, providing just 400 training and 100 testing images, which is extensively smaller than UCF-MultiOrgan-Path.

**Table 1.** This table provides a comparative analysis of existing histopathologic datasets, highlighting the newly introduced UCF-MultiOrgan-Path dataset, which consists of 977 whole slide images (WSIs) and approximately 2.38 million patches. The UCF-MultiOrgan-Path dataset enables comprehensive multi-organ classification, surpassing the capabilities of previously established specialized datasets.

| Dataset | Dataset Statistics | Dataset purpose |
| --- | --- | --- |
| Kimia Path24<sup>25</sup> | 24 WSIs, 24 different texture pattern | Benchmark dataset for the classification of 24 different texture patterns |
| Kimia Path24C<sup>26</sup> | Colored version of the Kimia Path24, same dataset statistics | Benchmark dataset for the classification of 24 different texture patterns |
| Atlas of Digital Pathology<sup>27</sup> | 100 WSIs, 17688 patches of 1088x1088 pixels | A generalized benchmark dataset for multi-label classification of histological tissue types from various organs with 57 distinct categories |
| CAMELYON16/17<sup>28–30</sup> | 399 WSIs (CAMELYON16) and 1,000 WSIs (CAMELYON17), 2 classes (metastasis or no metastasis in lymph nodes) | A benchmark dataset for binary classification of metastasis in lymph node tissue |
| TCGA<sup>31</sup> | Over 20,000 cancer tissue slides with over 30 cancer types across multiple organ | A comprehensive dataset primarily used for cancer classification across multiple organs |
| BACH<sup>32</sup> | 400 training, 100 testing microscopy images for four classes– normal, benign, in situ carcinoma, invasive carcinoma | A benchmark dataset for the classification of different breast cancer tissue types |
| PatchCamelyon<sup>33</sup> | 327,680 image patches, binary classification of Metastasis vs No Metastasis | A benchmark dataset for binary classification of tumor presence in lymph node patches |
| **UCF-MultiOrgan-Path** | **977 WSIs, approximately 2.38 million patches for 15 organ class** | **A benchmark dataset for organ classification** |

In summary, UCF-MultiOrgan-Path stands out due to its extensive dataset size and emphasis on multi-organ classification. It also has the potential to become a benchmark for developing and validating deep learning models across various tissue types, providing more versatility and generalizability than specialized datasets. By providing a diverse and annotated collection of whole slide histopathologic images aligned with FAIR principles, this publicly available dataset addresses a crucial need in digital pathology. Its realistic representation of clinical data makes it a valuable resource not only for organ classification but also for tackling more complex challenges. Furthermore, the extensive set of image patches enhances its utility for transfer learning, which is essential for refining models that can be applied in clinical settings. Overall, this dataset fosters the development of more robust deep-learning models that reflect clinical practice, thereby contributing to improved diagnostic accuracy and personalized treatment strategies.

## Methods

Our methodology encompassed a comprehensive dataset preparation process, carefully structured through several key steps of WSI pre-processing and patch pre-processing as illustrated in Figure 1. 1,700 tissue samples were collected through autopsy during the anatomy classes for students learning for 10 years (2010-2019) at the University of Central Florida (UCF). The UCF IRB issued a non-human subject determination for this study. The slides were stained with standard Hematoxylin and Eosin (H&E), Congo Red, GRAM, PAS-F, Trichrome, etc. The WSIs were prepared using an Aperio scanner at a magnification level of 20x. These WSIs represent a wide variety of tissues from various organs, ensuring a diverse and comprehensive dataset. We will discuss the technical validation in the subsequent section.

**Figure 1.**
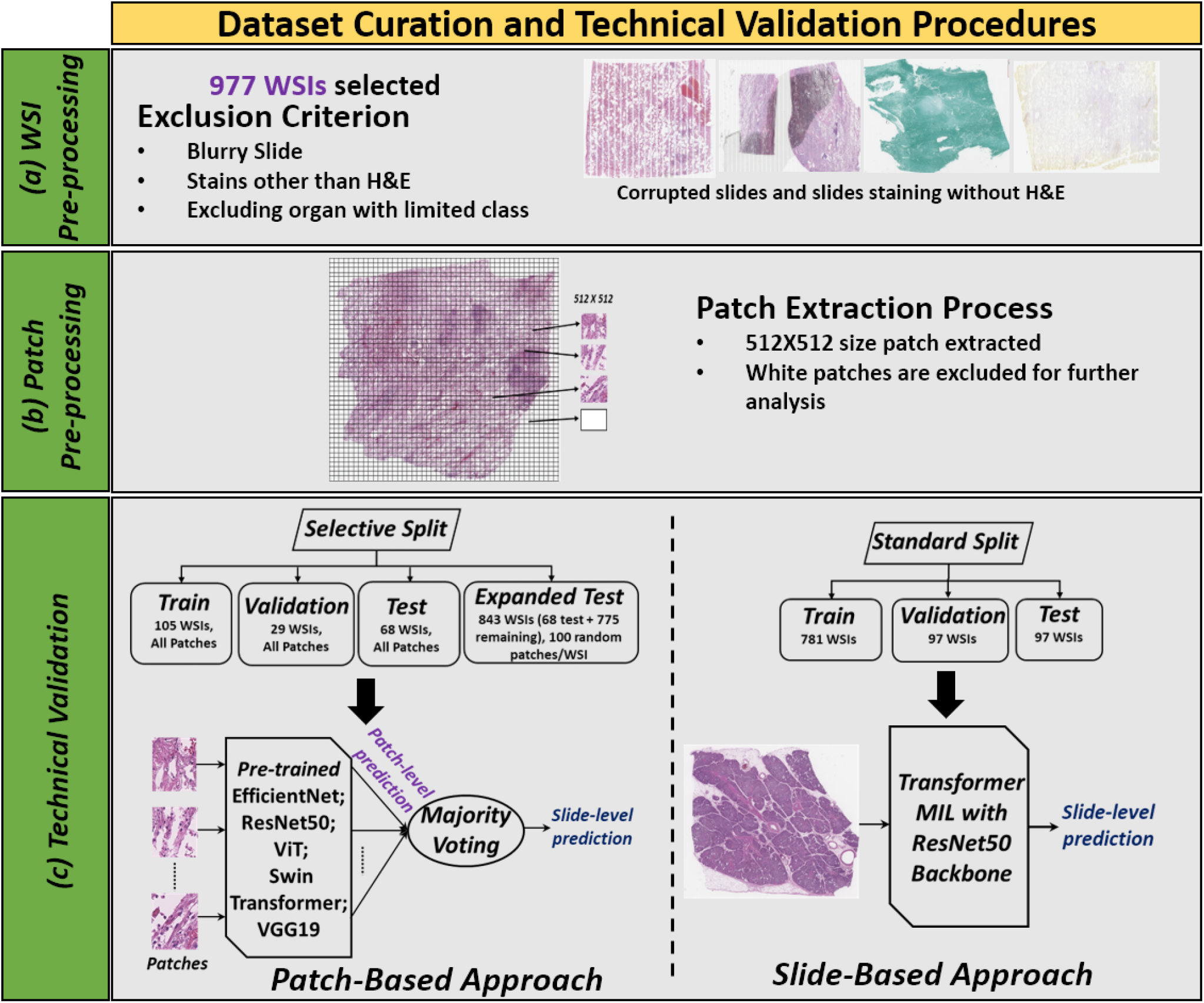
Overall process of dataset curation and validation procedures: (a) WSI Pre-processing: from the 1700 WSIs, we exclude slides that are blurry, limited class organs, and stains other than H&E to get 977 WSIs. (b) Patch Pre-processing: patches with a size of 512×512 are selected while excluding white spaces. (c) Two splits such as ***Selective and Standard Split*** are created to validate both patch and slide-based approaches, which are explained in detail in the Technical Validation section.

### WSI Pre-processing

We began by curating an extensive dataset of approximately 1,700 WSIs at 20x magnification derived from cadaver specimens with the assistance of three pathologists. These WSIs were purposefully selected to represent a wide range of tissues from various organs, ensuring a diverse and comprehensive dataset. To support machine learning analysis, we collaborated with Dr. Borowsky’s laboratory to digitize the WSIs, transforming them from their original wet slide format into digital pathology images. This digitization process enabled us to apply advanced deep-learning techniques to our research, enhancing our analytical capabilities. After the initial process of digitalization of 1700 WSIs, we selected 977 slides based on multiple criteria.

First, some slides became blurry or accumulated debris during the digitization process. To ensure a clean and robust dataset, we excluded these compromised slides from our analysis. The majority of slides are stained with H&E; therefore, due to the limited representation of organ classes in slides stained with other techniques and the variability introduced by different staining methods, we focused exclusively on slides with H&E staining. A sample image of corrupted and different WSI staining rather than H&E is presented in Figure S1 of the supplementary materials. Furthermore, certain organs such as the spinal cord, esophagus, and bone were represented by a very limited number of WSIs, complicating the division of data into training, testing, validation, and expanded test sets. Slides from these underrepresented organs were excluded from the final processed dataset. After applying all pre-processing steps, we finalized a dataset comprising 977 slides across 15 organ classes. A representative WSI image for each organ class is shown in Figure 2.

**Figure 2.**
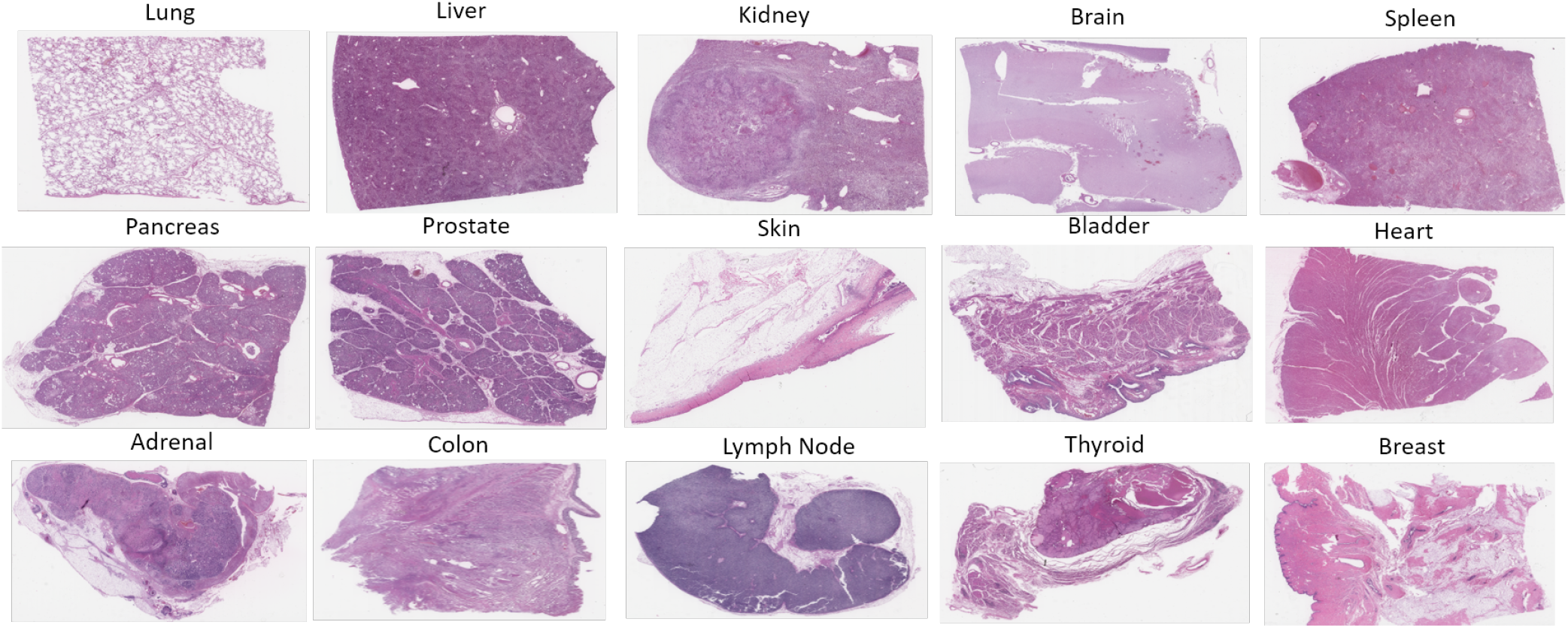
Sample WSI image for each organ class

### Patch Pre-processing

For patch extraction, we implemented a method previously described by Vrabac et al.^24^ to remove excessive white space from WSIs, ensuring that only relevant tissue areas are selected as patches. Non-overlapping patches were extracted at a resolution of 512×512 pixels to capture detailed patterns and textures within the tissue, while also maintaining computational efficiency to ensure compatibility with deep learning models^34,35^. White spaces were excluded through a two-step process. First, the RGB color space of each patch was converted to HSV color space, focusing on the saturation channel to differentiate between colored and white regions. Patches were excluded if the proportion of pixels with low saturation exceeded a predefined threshold, indicating a predominance of white space. In the second step, patches were converted to grayscale, and the Sobel operator was applied to detect edges and compute gradient magnitude. Patches with a high proportion of pixels having zero gradient magnitude, which also indicates white space, were excluded from further analysis. This combination of techniques allowed us to effectively eliminate patches with excessive white space, ensuring that the retained patches were informative and suitable for subsequent analysis. Multiple patches for each organ class are provided in the supplementary materials.

### Data Records

The WSIs included in this dataset are accessible at https://stars.library.ucf.edu/ucfnecropsywsi/. The WSI images have been uploaded as bundles of approximately 30 WSIs per bundle. Each dataset bundle is designated as UCF WSI Batch XX, with specific sample bundles referenced in^36,37^. In addition to the WSIs, 2,379,949 image patches from 15 organ classes have been included on the same website. The patch files have been organized and bundled by anatomical region, with each zip file approximately 20 GB in size to facilitate usability and ease of downloading. The number of patches per zip file and the number of zip files for each organ vary depending on the dataset. Each zip file is clearly labeled by organ type and batch number (e.g., UCF Adrenal Patch, UCF Bladder Patch Batch 01, UCF Brain Patch Batch 03)^38–52^, enabling users to efficiently locate and use the data for their research needs. For larger zip files exceeding 20 GB, the patch data has been split into multiple parts using a split archive system for ease of use, with examples including UCF Heart Patch Batch, UCF Heart Patch Batch 01, UCF Heart Patch Batch 02, …, up to UCF Heart Patch Batch 09. To recombine these files, ensure all the split parts are present in the same directory, use a compatible unarchiving tool like WinRAR or 7-Zip, open the main .zip file (e.g., UCF Heart Patch Batch), and extract the data following the software instructions. Overall, the site hosts over 2 TB of compressed zip data, providing a comprehensive and valuable resource for researchers.

### Technical Validation

We conduct a comprehensive technical validation of the dataset and explore the effectiveness of state-of-the-art (SOTA) deep learning models on the UCF-MultiOrgan-Path dataset both at the patch and slide level classification. To achieve this, we provide two approaches such as patch-based approach and a slide-based approach, as illustrated in Figure 1. In the patch-based approach, individual patches are input into the deep learning model for classification, with the final slide classification determined by majority voting across the patch predictions. In the slide-based approach, MIL^58^ is used to classify the slide. Both the patch and slide-based methods are used due to their prevalence and to provide different aspects of analysis of the data.

### Patch-based Approach

#### Dataset Splitting

Whole slide images are split into distinct sets for training, validation, testing, and expanded testing, which we refer to as the ***Selective Split***. Specifically, we randomly selected 105 WSIs for the training set, 29 for the validation set, and 68 for the test set. The remaining 775 WSIs were combined with the initial test set to form an expanded test set, totaling 843 WSIs. The total number of WSIs and patches for each organ class in each set, along with the overall slide and patch counts for each organ, is presented in Table **4**.

The primary goal of this study is to introduce the dataset and provide initial validation, as training and validating a deep learning model on the entire dataset of nearly 2.38 million patches would be time-consuming. Therefore, we limited our analysis to the aforementioned number of slides. To expedite the validation process and reduce computational overhead, we performed a selective evaluation by randomly sampling 100 patches from the WSIs in the expanded test set. This ***Selective Split*** approach enables an efficient and representative assessment of the model’s performance without the need to use the entire patch set for each WSI. We encourage other researchers to conduct experiments using the complete dataset to facilitate the development of pre-trained models for pathology patches. These models could serve as valuable pre-trained encoders for transfer learning applications, such as classification or segmentation tasks in digital pathology.

#### Implementation Details

Five pre-trained backbones—EfficientNet^53^, ResNet50^54^, Vision Transformer (ViT)^55^, Swin Transformer^56^, and VGG19^57^—were selected to evaluate our dataset’s performance. These architectures represent a mix of both convolutional and transformer-based architectures, allowing a robust evaluation of the dataset’s performance across diverse model types.

All the patches extracted from the train and validation set (Figure 1) are used for model training and validation. All models included in this study are trained using the PyTorch framework, employing a Tesla T4 GPU (NVIDIA, Santa Clara, CA). The method is trained to perform patch-level classification and to achieve slide-level classification through majority voting. As the primary objective of this paper is to introduce an organ classification method and a single epoch takes a lot of time to run, we trained all the models with 5 epochs only. The patches are resized to 224×224 pixels to be compatible with the deep learning models. A batch size of 128, Adam as the optimizer^59^, and cross-entropy loss function are used to train the models. To validate the trained models, we utilize both the test and expanded test set and provide the results as patch-level and slide-level predictions in Table 2 and 3. Accuracy, precision, recall, and F1-Score are used as the primary evaluation metrics to assess the model’s performance, providing a comprehensive view of both classification correctness and the balance between false positives and false negatives.

**Table 2.** Accuracy, precision, recall, F1-Score of different deep-learning models for patch and slide level classification task for **test set** for patch-based approach for ***Selective Split***.

| Model | Patch Level |  |  |  | Slide Level |  |  |  |
| --- | --- | --- | --- | --- | --- | --- | --- | --- |
|  | Accuracy | Precision | Recall | F1-Score | Accuracy | Precision | Recall | F1-Score |
| EfficientNet<sup>53</sup> | 60.20 | 60.28 | 60.20 | 58.74 | 69.12 | 77.37 | 69.12 | 67.28 |
| ResNet50<sup>54</sup> | 59.82 | 59.17 | 59.82 | 57.96 | 69.12 | 74.98 | 69.12 | 65.92 |
| ViT<sup>55</sup> | 60.02 | 58.91 | 60.02 | 58.61 | 76.47 | 78.08 | 76.47 | 75.35 |
| Swin Transformer<sup>56</sup> | 62.11 | 62.21 | 62.11 | 60.55 | 73.53 | 79.75 | 73.53 | 72.22 |
| VGG19<sup>57</sup> | 57.91 | 57.28 | 57.91 | 54.97 | 63.24 | 65.72 | 63.24 | 57.95 |

**Table 3.** Accuracy, precision, recall, F1-Score of different deep-learning models for patch and slide level classification task for **expanded test set** for patch-based approach for ***Selective Split***.

| Model | Patch-level |  |  |  | Slide-level |  |  |  |
| --- | --- | --- | --- | --- | --- | --- | --- | --- |
|  | Accuracy | Precision | Recall | F1-Score | Accuracy | Precision | Recall | F1-Score |
| EfficientNet<sup>53</sup> | 48.05 | 73.66 | 48.05 | 53.41 | 67.62 | 84.86 | 67.62 | 72.26 |
| ResNet50<sup>54</sup> | 46.69 | 72.47 | 46.69 | 51.32 | 63.70 | 81.23 | 63.70 | 67.17 |
| ViT<sup>55</sup> | 46.05 | 72.77 | 46.05 | 49.95 | 60.85 | 81.64 | 60.85 | 64.58 |
| Swin Transformer<sup>56</sup> | 47.92 | 73.75 | 47.92 | 52.65 | 64.65 | 83.95 | 64.65 | 68.80 |
| VGG19<sup>57</sup> | 41.83 | 70.05 | 41.83 | 42.84 | 54.69 | 79.53 | 54.69 | 55.24 |

#### Results and Discussion

The patch-based approach provides notable variability in performance across different deep learning models, with Swin Transformer achieving the highest accuracy of 62.11% for patch-level prediction on the test set and ViT with an accuracy of 76.47% for slide-level prediction. However, both the patch-level and slide-level accuracy decreased for the expanded test set due to the use of 100 randomly sampled patches. Random sampling of fewer patches increases the likelihood of missing distinguishing features or selecting less informative tissues with more background and less distinctive patterns, creating sampling bias and potentially reducing prediction accuracy.

In Figure 3, 4, 5, 6, we present confusion matrices for patch-level and slide-level prediction for both test and expanded test sets using ViT to illustrate the accuracy of each organ class separately. Additionally, confusion matrices and classification results for each organ for all the models are provided in the supplementary materials. Sample patches for all the organs are also provided in the supplementary materials. This class-wise analysis reveals that organs such as the brain and heart, which possess distinctive histopathological features, consistently achieved higher F1-Scores indicating fewer false positives and false negatives. On the other hand, lower accuracy of lung in the test set may be due to the limited number of patches (4,013) derived from only five test slides, as shown in Table 4. This limited patch counts likely results in fewer informative patches, which may not fully capture the histological diversity needed for accurate classification. Increasing the number of lung slides in the training set could help the model learn to classify lung tissue more accurately. The lower accuracy for adrenal and colon classification may also be due to the relatively small number of slides and patches used for training these organs compared to others (Table 4). With fewer examples, the model may struggle to generalize well for these tissues. Additionally, confusion matrices for most models reveal that lymph nodes are frequently misclassified as the spleen, as these tissues have similar histological features such as lymphoid follicles (Figure S6, S14) making it challenging for the model to distinguish them accurately. Interestingly, lymph nodes are often misclassified as spleen, but the opposite is less common. This could be due to patch distribution, as shown in Table 4, where the spleen has approximately 1.5 times more training patches than the lymph node. This imbalance may cause the model to be more biased toward classifying lymph node patches as the spleen.

**Table 4.** Number of slides and patches used in train, validation, test, and expanded test set in ***Selective Split*** for model training and validation, along with total slide and patch count for each organ.

| Organ Name | Slides |  |  |  |  | Patches |  |  |  |  |
| --- | --- | --- | --- | --- | --- | --- | --- | --- | --- | --- |
|  | Train | Val | Test | Exp. Test | Total | Train | Val | Test | Exp. Test | Total |
| Lung | 8 | 2 | 5 | 374 | 384 | 17856 | 1533 | 4013 | 37400 | 627760 |
| Liver | 8 | 2 | 5 | 79 | 89 | 29546 | 7209 | 21435 | 7900 | 336676 |
| Kidney | 8 | 2 | 5 | 82 | 92 | 22858 | 7949 | 16074 | 8200 | 286876 |
| Brain | 8 | 2 | 5 | 29 | 39 | 38534 | 9803 | 18933 | 2900 | 161173 |
| Spleen | 8 | 2 | 5 | 14 | 24 | 31848 | 7223 | 19231 | 1400 | 87053 |
| Pancreas | 8 | 2 | 5 | 18 | 28 | 19839 | 4502 | 10772 | 1800 | 67053 |
| Prostate | 8 | 2 | 5 | 47 | 57 | 25455 | 7097 | 19792 | 4700 | 205439 |
| Skin | 8 | 2 | 5 | 57 | 67 | 2406 | 1223 | 1536 | 5323 | 32118 |
| Bladder | 8 | 2 | 5 | 19 | 29 | 16952 | 4502 | 12852 | 1900 | 61474 |
| Heart | 8 | 2 | 5 | 102 | 112 | 27267 | 7739 | 17250 | 10200 | 372844 |
| Adrenal | 4 | 2 | 3 | 3 | 9 | 13530 | 5945 | 8642 | 300 | 28117 |
| Colon | 3 | 1 | 3 | 3 | 7 | 10179 | 1701 | 8676 | 300 | 20556 |
| Lymph Node | 8 | 2 | 5 | 9 | 19 | 21864 | 6288 | 13205 | 900 | 50008 |
| Thyroid | 5 | 2 | 3 | 3 | 10 | 12248 | 1685 | 4951 | 300 | 18884 |
| Breast | 5 | 2 | 4 | 4 | 11 | 10994 | 5400 | 7524 | 400 | 23918 |
| Total | 105 | 29 | 68 | 843 | 977 | 301376 | 79799 | 184886 | 83923 | 2379949 |

**Figure 3.**
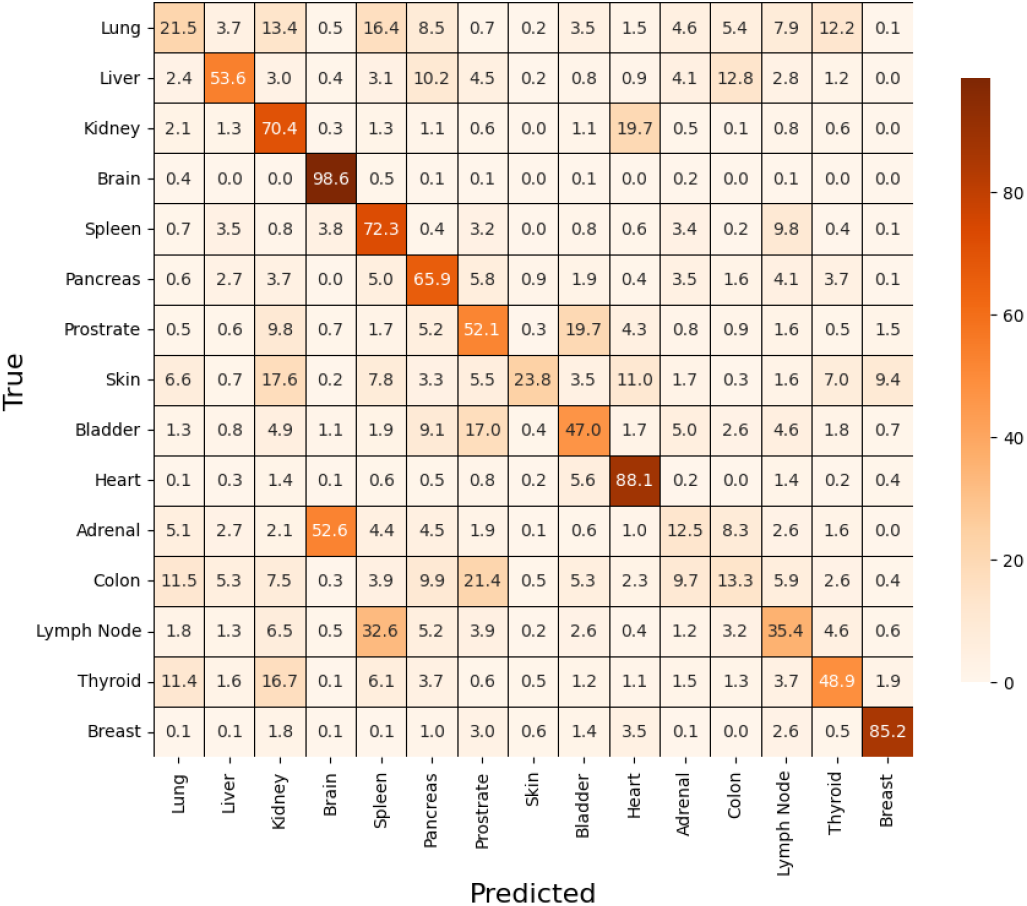
Confusion matrix for patch-level classification for test set using **ViT**

**Figure 4.**
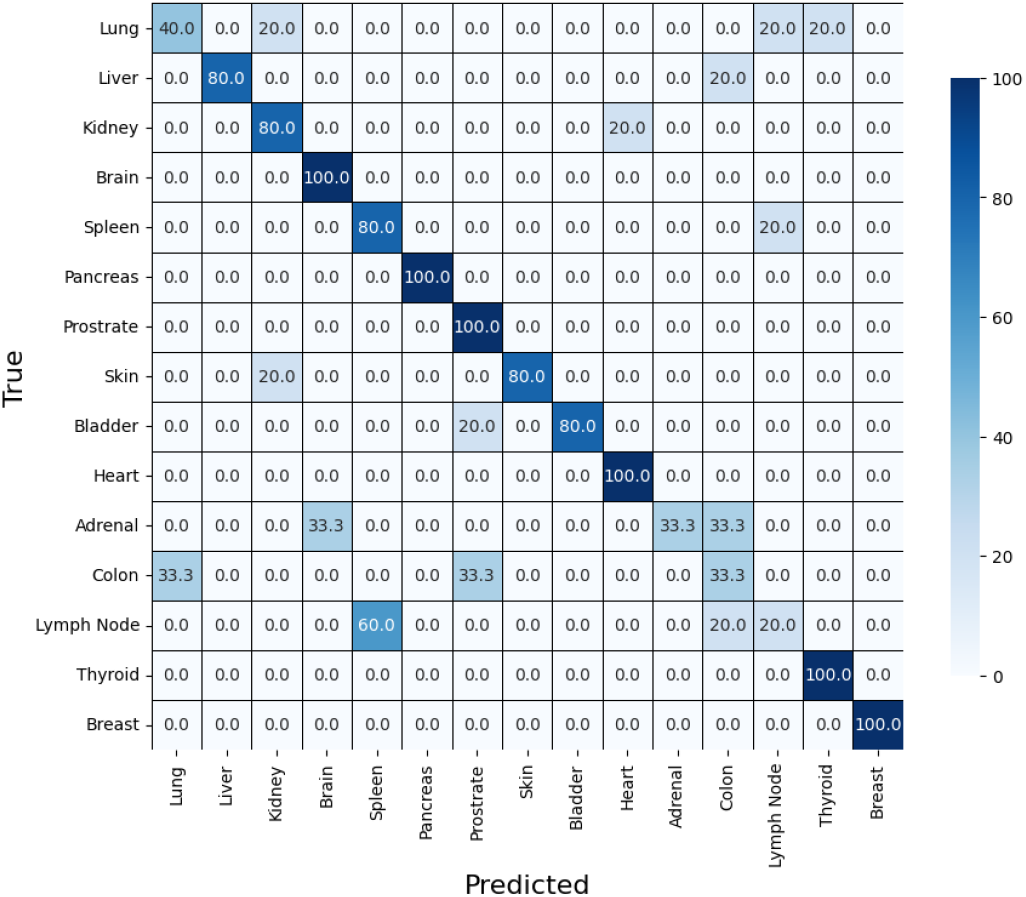
Confusion matrix for slide-level classification for test set using **ViT**

**Figure 5.**
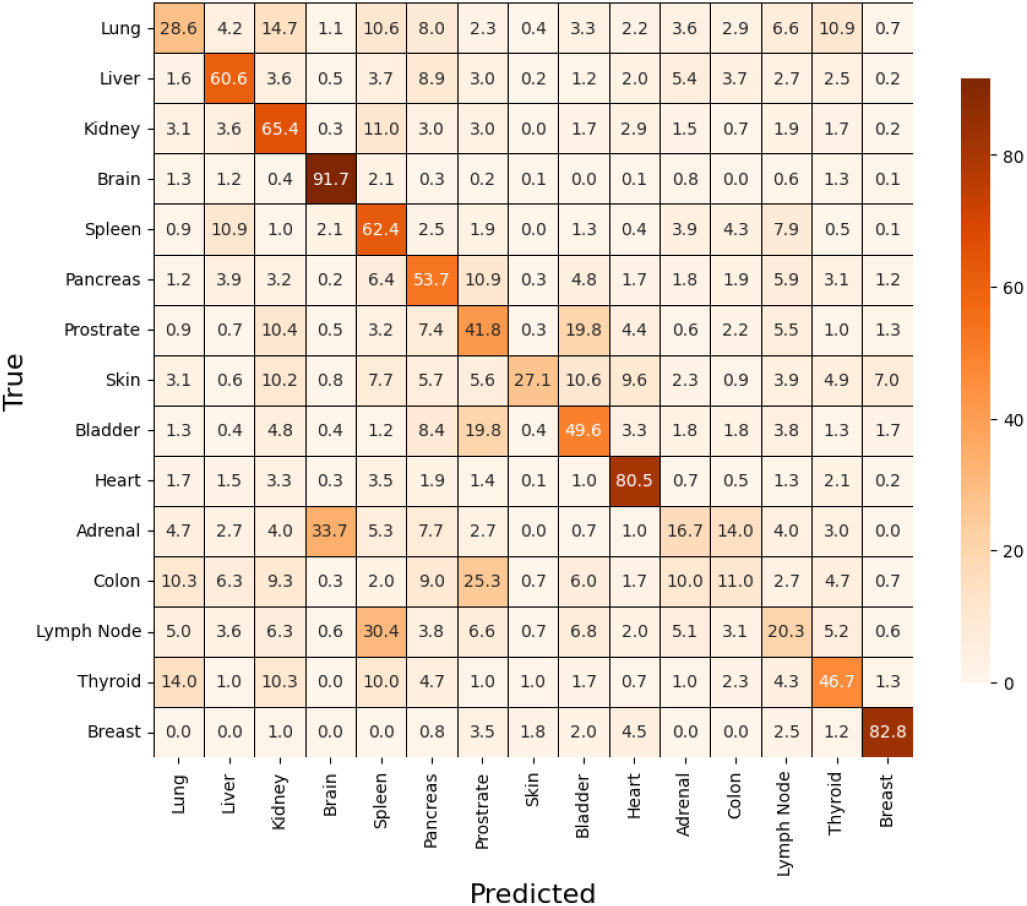
Confusion matrix for patch-level classification for expanded test set using **ViT**

**Figure 6.**
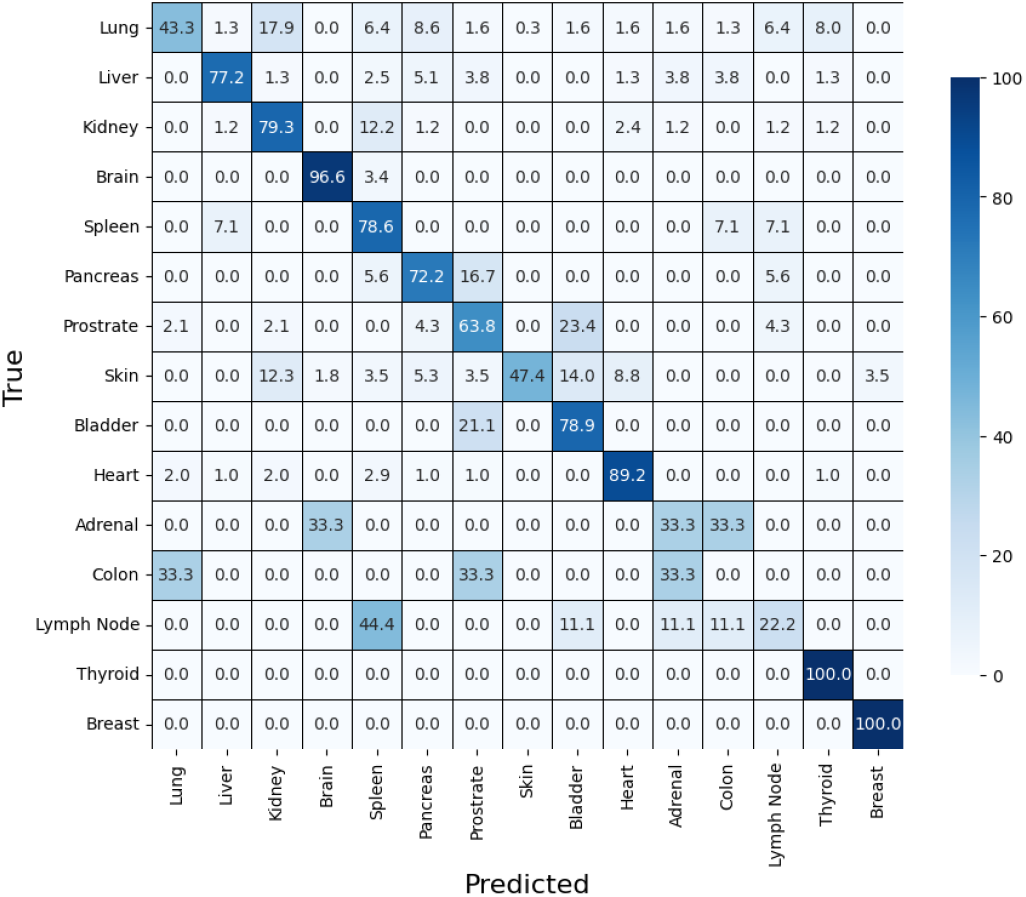
Confusion matrix for slide-level classification for expanded test set using **ViT**

The confusion matrices highlight common misclassification patterns, particularly among organs with similar structures, suggesting the need for balanced datasets and advanced sampling techniques. Additionally, as we only experimented with a patch size of 512×512 pixels, future work could explore using patches of varying sizes to assess how different scales impact classification accuracy for the patch-based approach. Overall, the findings highlight the ability of various models to capture complex patterns for multi-organ classification, while also pointing to areas for future improvement in addressing class imbalance and optimizing patch selection strategies.

### Slide-based Approach

#### Dataset Splitting

We use a different splitting scheme, called the ***Standard Split***, which applies a random 80%-10%-10% train-validation-test split to the 977 WSIs, in contrast to the ***Selective Split***. This approach results in 781 WSIs for the training set, 97 WSIs for the validation set, and 99 WSIs for the test set. This splitting scheme allows for consistent dataset evaluation by leveraging the MIL approach to utilize the entire dataset efficiently. Unlike the patch-based method, it minimizes training complexity regarding time and computational resources.

#### Implementation Details

MIL is a weakly-supervised deep learning approach where a single class label is assigned to a bag of instances^58^. Transformer MIL^60^, with a ResNet50^54^ backbone, is the improved version of the original MIL paper chosen as the slide-based method for evaluating the UCF-MultiOrgan-Path dataset. For the sake of simplicity, we refer to Transformer MIL as MIL in this paper. The approach selects random patches from each WSI, trains a backbone to represent each patch as a feature, and then uses an attention and multi-layer perception head for classification.

Patches of size 256x256 are chosen from each WSI and undergo random flip and rotation augmentations during training. The AdamW optimizer, binary cross-entropy loss, and a cosine annealing scheduler are adapted from the original implementation. Unlike the original paper, we only select 30 random patches from each WSI rather than 56. The slide-based approach used the same hardware and platform as the patch-based approach.

#### Results and Discussion

In the ***Standard Split*** where the classes are balanced for each split. This split focuses more on evaluating the whole dataset, where the MIL method achieves 55.45% accuracy, shown in Table 5. We observe in Figure 7 that due to the class imbalance, MIL achieves 84.6% on lung classification and 0.0% accuracy on seven of the other organs. Notably, the model manages to accurately (100%) predict the brain organ even with only 39 slides total which, suggests how starkly different the cellular structure of the brain is compared to other organ classes (Figure S5). These results illustrate the rigor of the multi-organ classification task with slide-level approaches and the need for additional datasets in this direction. Additionally, we managed to achieve a 64.54% accuracy in Table 5 with the MIL method when expanding the random patch selection count from 30 to 44. With additional computational resources, the MIL method would be able to achieve even higher accuracy; however, it would still not be as effective as a patch-based approach simply due to the difference in granularity of annotations. The confusion matrix for a patch count of 44 is provided in Figure 8 to illustrate how accurately each specific organ is classified using this approach.

**Table 5.** Precision, Recall, F1-Score for each organ class on ***Standard Split*** using **MIL** with Patch Count 30 and 44.

| Organ | Patch Count = 30 |  |  | Patch Count = 44 |  |  |
| --- | --- | --- | --- | --- | --- | --- |
|  | Precision | Recall | F1-Score | Precision | Recall | F1-Score |
| Lung | 84.62 | 84.62 | 84.62 | 94.44 | 87.18 | 90.67 |
| Liver | 28.57 | 20.00 | 23.53 | 80.00 | 80.00 | 80.00 |
| Kidney | 69.23 | 90.00 | 78.26 | 64.29 | 90.00 | 75.00 |
| Brain | 45.45 | 100.00 | 62.50 | 66.67 | 80.00 | 72.73 |
| Spleen | 0.00 | 0.00 | 0.00 | 100.00 | 33.33 | 50.00 |
| Pancreas | 0.00 | 0.00 | 0.00 | 0.00 | 0.00 | 0.00 |
| Prostrate | 0.00 | 0.00 | 0.00 | 50.00 | 28.57 | 36.36 |
| Skin | 35.00 | 87.50 | 50.00 | 28.57 | 100.00 | 44.44 |
| Bladder | 12.50 | 25.00 | 16.67 | 0.00 | 0.00 | 0.00 |
| Heart | 60.00 | 25.00 | 35.29 | 100.00 | 41.67 | 58.82 |
| Adrenal | 0.00 | 0.00 | 0.00 | 0.00 | 0.00 | 0.00 |
| Colon | 0.00 | 0.00 | 0.00 | 0.00 | 0.00 | 0.00 |
| Lymph Node | 0.00 | 0.00 | 0.00 | 0.00 | 0.00 | 0.00 |
| Thyroid | 100.00 | 100.00 | 100.00 | 0.00 | 0.00 | 0.00 |
| Breast | 0.00 | 0.00 | 0.00 | 0.00 | 0.00 | 0.00 |
| **Accuracy** | 55.45 |  |  | 64.54 |  |  |

**Figure 7.**
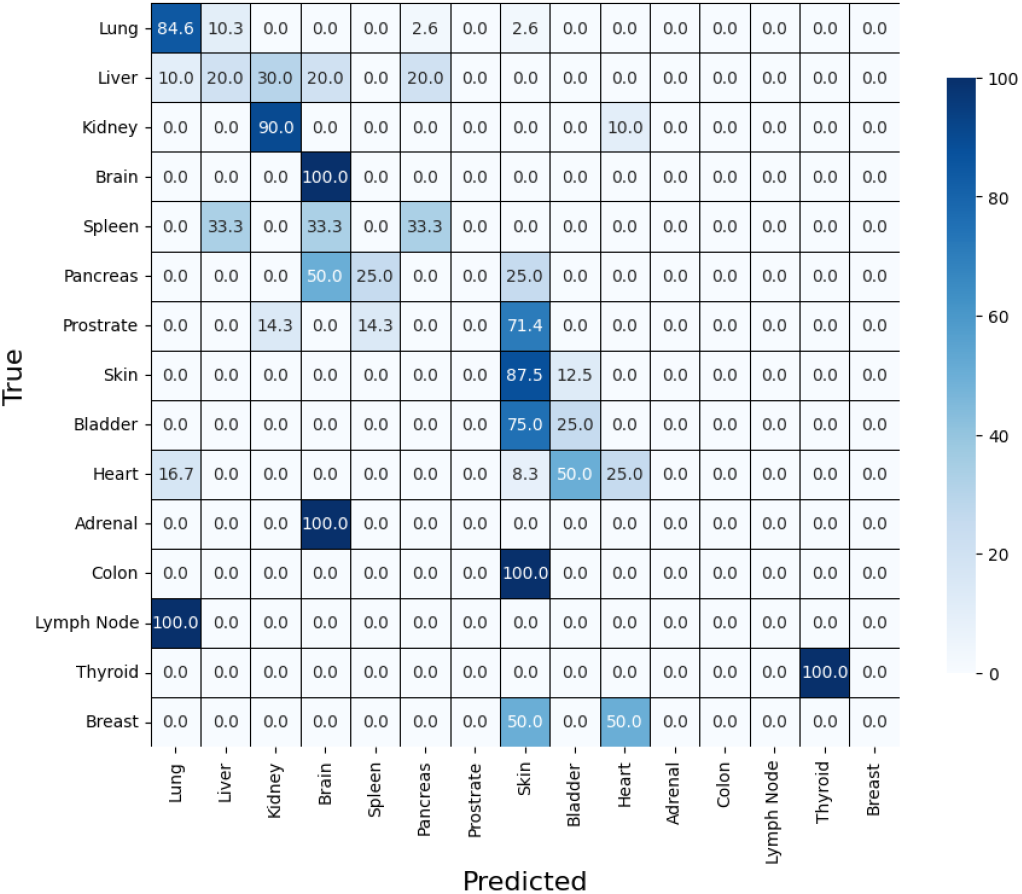
Confusion matrix on ***Standard Split*** using **MIL** with Patch Count equal to 30

**Figure 8.**
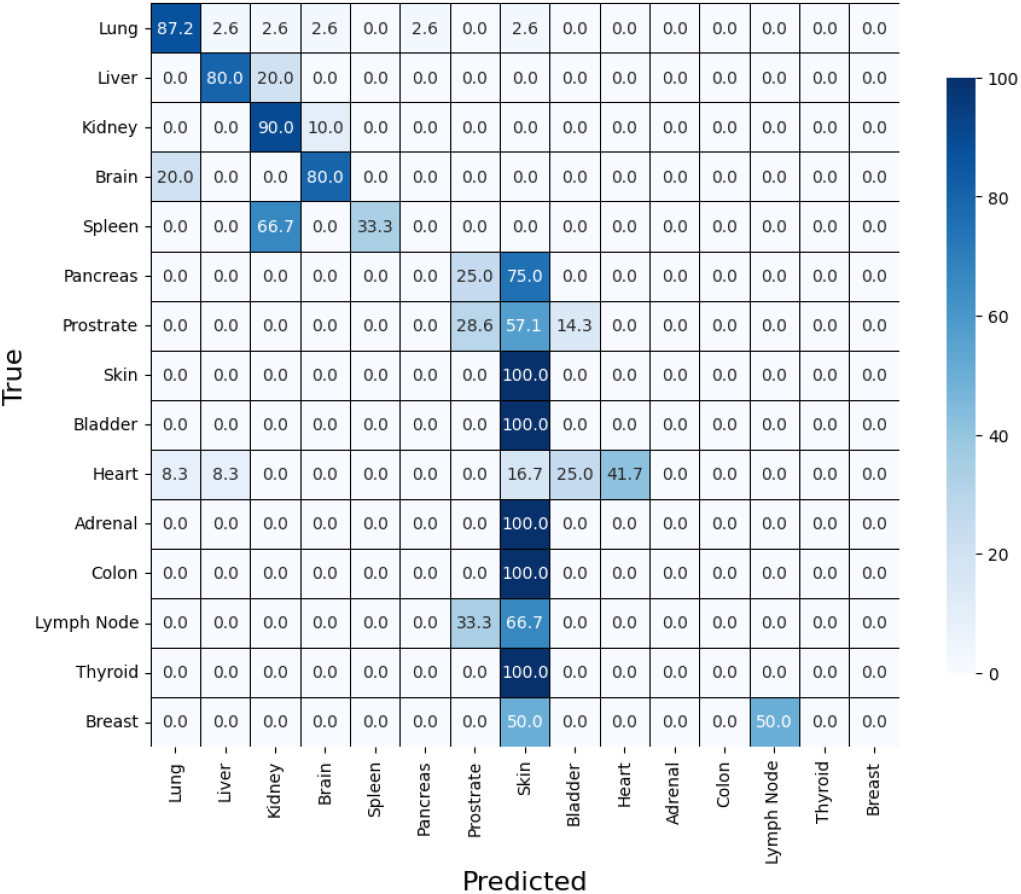
Confusion matrix on ***Standard Split*** with Patch Count equal to 44

Future work could focus on developing an adaptive patch selection approach to identify informative patches unique to specific organ classes, thereby enhancing classification accuracy. Increasing the number of patches selected per WSI could further improve the model’s ability to capture diverse tissue characteristics; however, this approach would require substantial GPU memory and computational resources.

In conclusion, the potential of UCF-MultiOrgan-Path lies in its ability to address the limitations of existing datasets, such as limited organ diversity, small patch counts, and narrow focus on specific diseases or textures. By offering a wide range of organ types, this dataset enables the development and validation of deep learning models that can learn richer, more diverse features, resulting in robust, generalizable models suited for real-world clinical applications. With approximately 2.38 million patches, UCF-MultiOrgan-Path allows models to capture complex histological variations and subtle patterns that smaller datasets often miss. This comprehensive, large-scale resource fills a critical gap in histopathologic research, supporting the development of more accurate and scalable models. While the dataset’s size presents computational challenges, requiring powerful resources such as GPUs with high memory and extended training time, it also provides a valuable benchmark for advancing computational pathology and clinical diagnostic tools, bridging the gap between academic research and practical applications.

## Supporting information

Additional Results and Figures

## Data Availability

The WSIs included in this dataset are accessible at https://stars.library.ucf.edu/ucfnecropsywsi/.

## Usage Notes

The dataset is the property of the University of Central Florida, which holds all rights to it. Licensors provide non-exclusive rights to utilize the dataset for research purposes, free of charge, to both academic and industrial research users. However, sublicensing rights are not granted. Usage is limited to non-commercial purposes, specifically for research and/or evaluation only. Subject to the terms and conditions of this License, users are granted a non-exclusive, royalty-free license to reproduce, prepare derivative works of, publicly display, publicly perform, and distribute the dataset and any resulting derivative works in any form.

## Code availability

The code for patch extraction, models of technical validation, validation results, and patch distribution for training are publicly accessible on GitHub. Visit our GitHub repository at https://github.com/Md-Sanzid-Bin-Hossain/UCF-WSI-Dataset to explore and contribute to our work.

## Author contributions statement

Md.H. performed technical validation, data pre-processing, analysis, and contributed to manuscript writing. Y.P. conducted data annotation. J.B. provided background research, performed technical validation, prepared data documentation, and contributed to manuscript writing. A.B. performed technical validation and contributed to manuscript writing. Mi.H. provided background research and contributed to manuscript writing. S.F. performed data pre-processing and data documentation. A.B. performed digitization of WSIs. H.K. helped with the data annotation. C.R. provided background research. A.F. helped with data collection. B.W. helped with data collection C.C. performed technical validation. L.W. performed technical validation. M.H. collected and annotated data. D.H. conceived the project, provided background research, performed data pre-processing and analysis, and conducted technical validation. All authors reviewed the manuscript.

