## Additional Results and Figures for "UCF-MultiOrgan-Path:A Benchmark Dataset of Histopathologic Images for Deep Learning-Based Organ Classification"

### Supplementary Materials

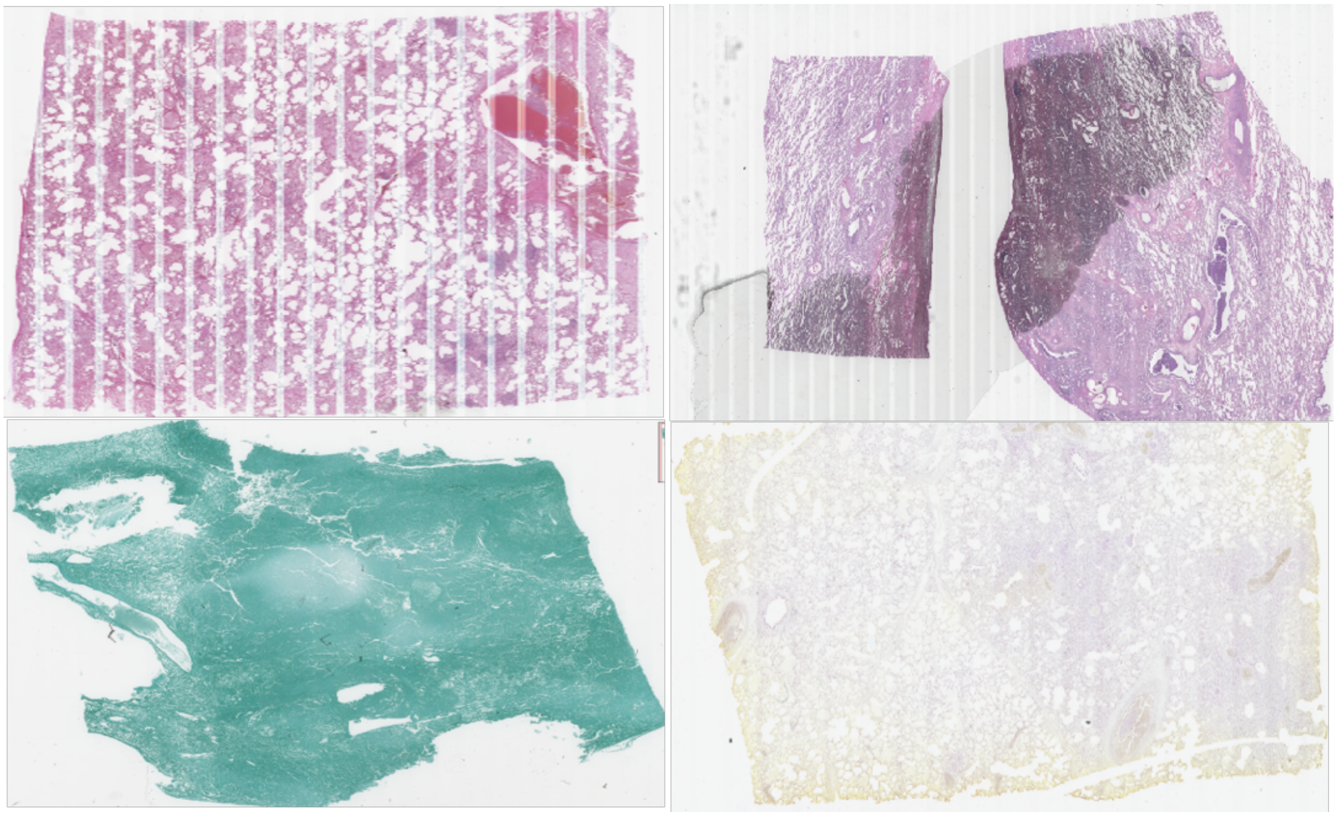

**Figure S1.** Corrupted and slides with different staining techniques other than H&E

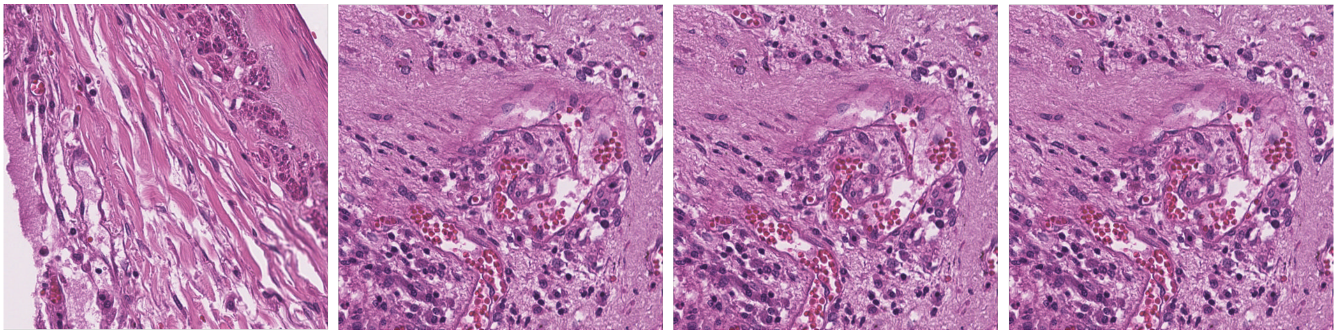

**Figure S2.** Example of Lung patches

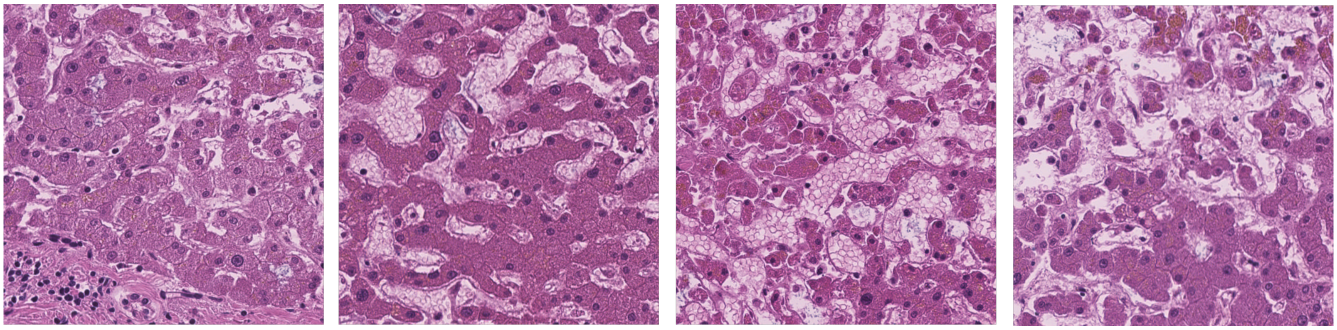

**Figure S3.** Example of Liver patches

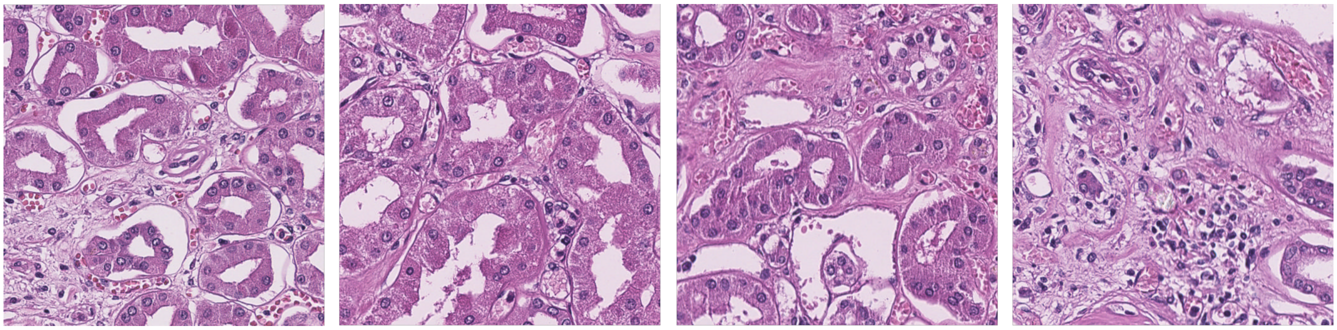

**Figure S4.** Example of Kidney patches

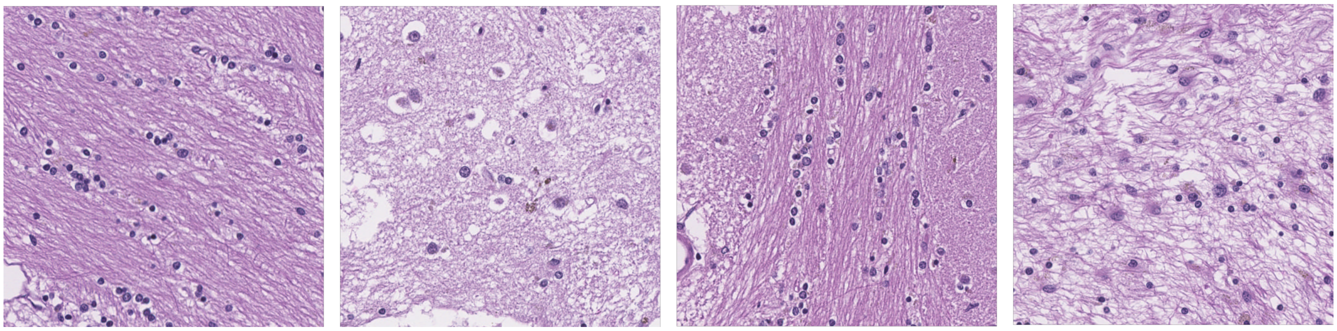

**Figure S5.** Example of Brain patches

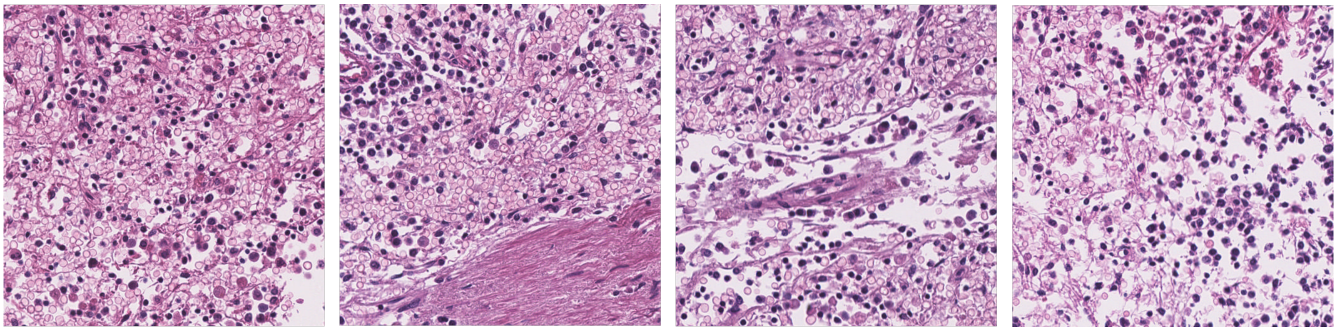

**Figure S6.** Example of Spleen patches

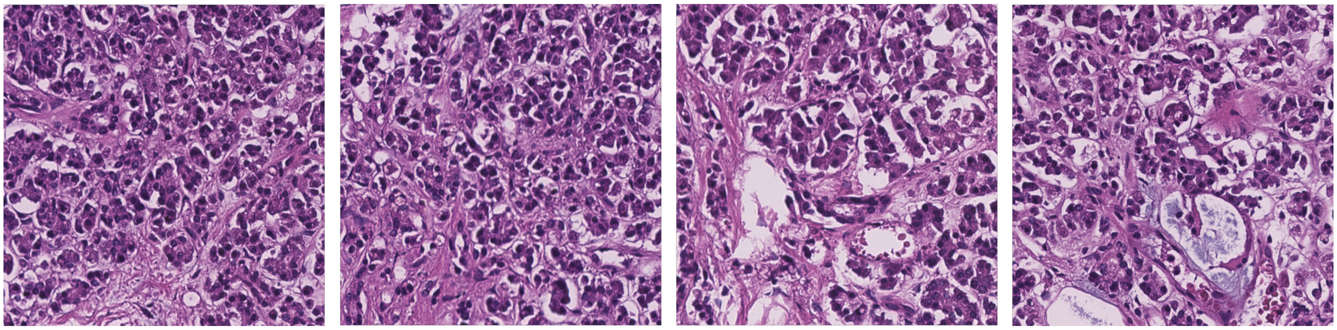

**Figure S7.** Example of Pancreas patches

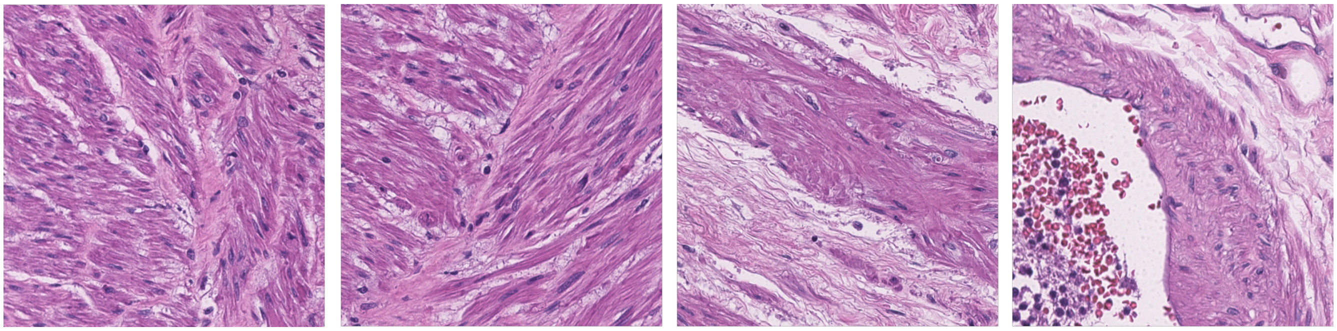

**Figure S8.** Example of Prostate patches

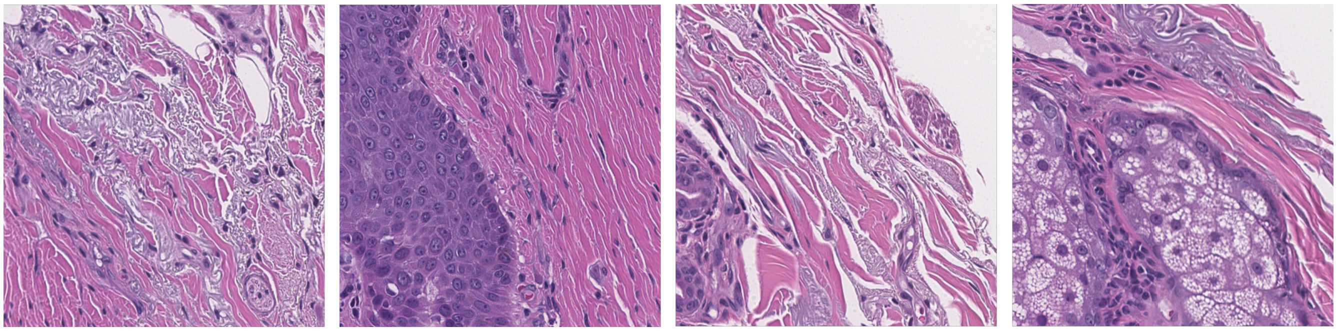

**Figure S9.** Example of Skin patches

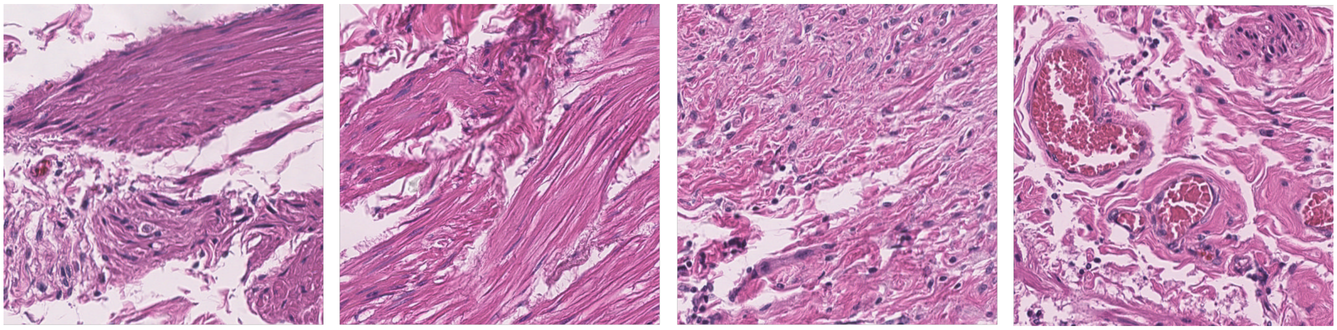

**Figure S10.** Example of Bladder patches

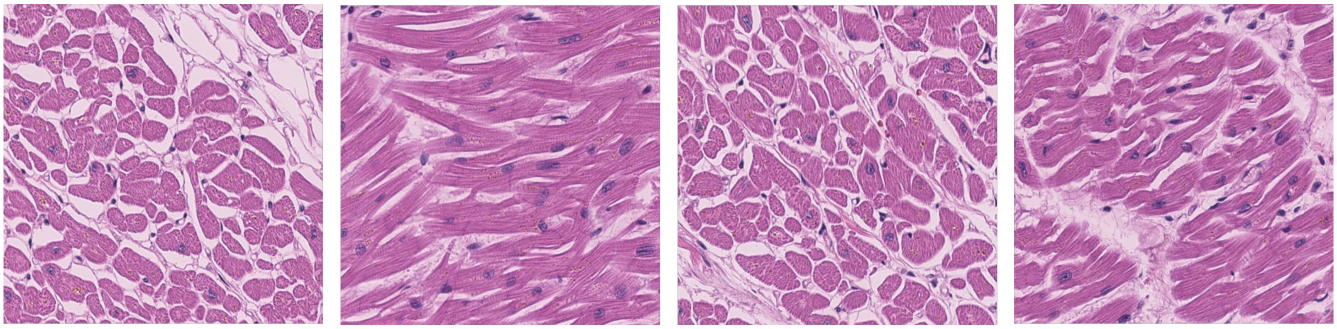

**Figure S11.** Example of Heart patches

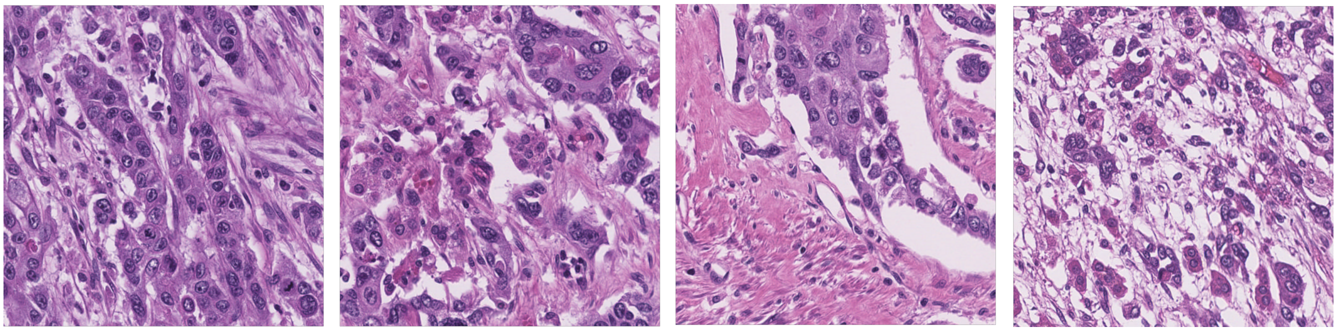

**Figure S12.** Example of Adrenal patches

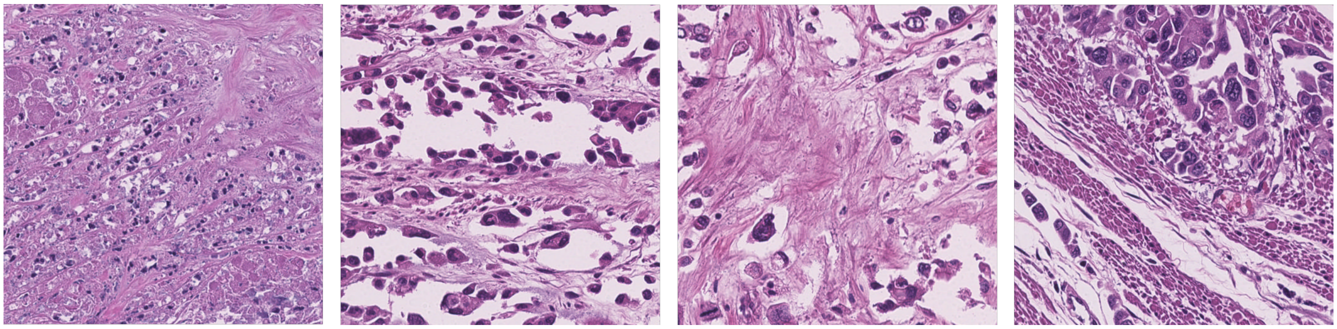

**Figure S13.** Example of Colon patches

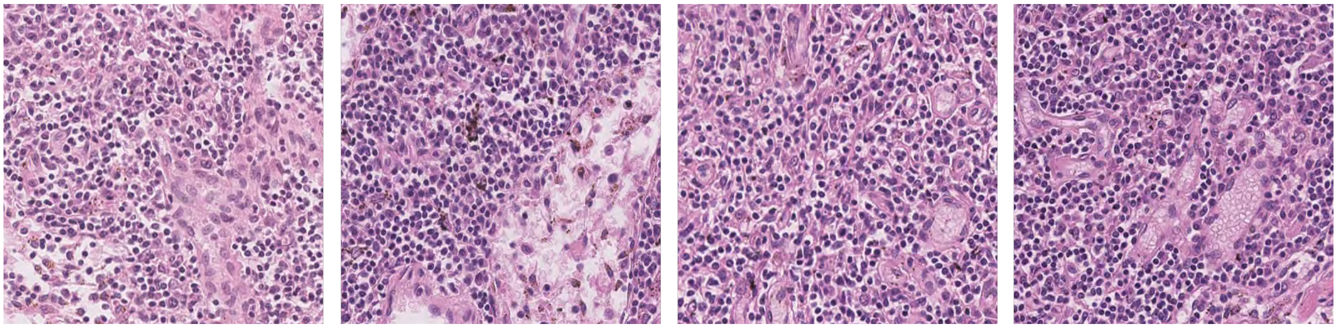

**Figure S14.** Example of Lymph Node patches

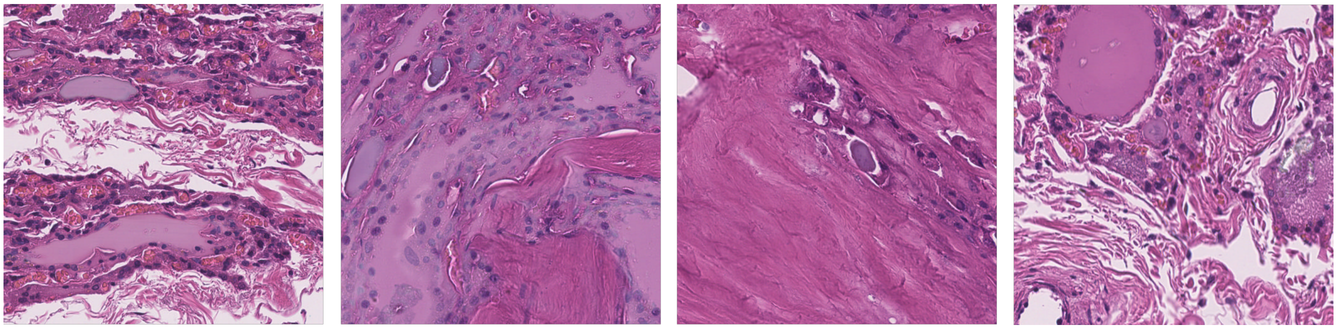

**Figure S15.** Example of Thyroid patches

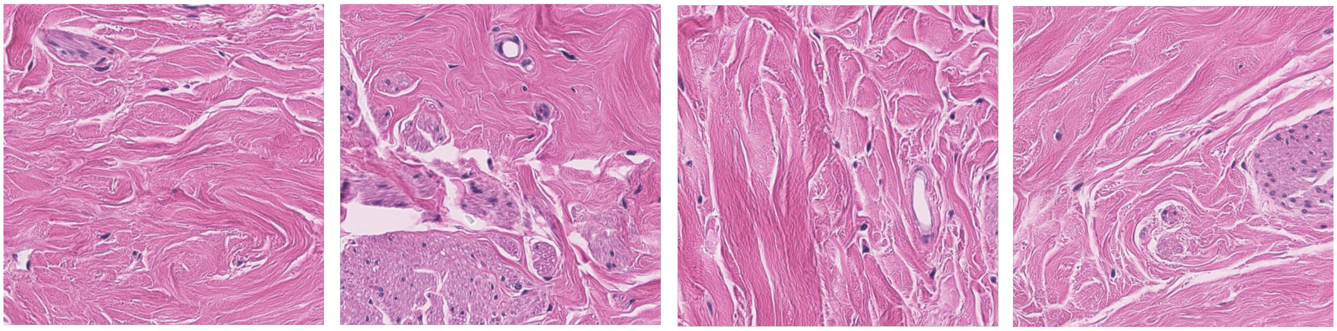

**Figure S16.** Example of Breast patches

**Table S1.** Precision, Recall, F1-Score for each organ class separately for test and expanded test set for both patch and slide-level classification using **EfficientNet**

| Organ | Test | | | | | | Expanded Test | | | | | |
|---|---|---|---|---|---|---|---|---|---|---|---|---|
| | patch-level | | | slide-level | | | patch-level | | | slide-level | | |
| | Precision | Recall | F1-Score | Precision | Recall | F1-Score | Precision | Recall | F1-Score | Precision | Recall | F1-Score |
| Lung | 24.78 | 35.14 | 29.07 | 100.00 | 20.00 | 33.33 | 90.06 | 34.90 | 50.30 | 94.80 | 63.37 | 75.96 |
| Liver | 82.18 | 46.23 | 59.17 | 100.00 | 60.00 | 75.00 | 73.45 | 53.10 | 61.64 | 85.07 | 72.15 | 78.08 |
| Kidney | 73.39 | 62.12 | 67.28 | 80.00 | 80.00 | 80.00 | 56.17 | 60.48 | 58.25 | 79.10 | 64.63 | 71.14 |
| Brain | 77.83 | 97.45 | 86.55 | 83.33 | 100.00 | 90.91 | 77.26 | 90.66 | 83.42 | 90.32 | 96.55 | 93.33 |
| Spleen | 72.62 | 78.22 | 75.32 | 71.43 | 100.00 | 83.33 | 17.62 | 66.50 | 27.86 | 22.45 | 78.57 | 34.92 |
| Pancreas | 43.97 | 73.64 | 55.06 | 71.43 | 100.00 | 83.33 | 14.54 | 57.83 | 23.23 | 20.29 | 77.78 | 32.18 |
| Prostrate | 48.67 | 54.19 | 51.28 | 37.50 | 60.00 | 46.15 | 36.26 | 45.21 | 40.25 | 50.00 | 51.06 | 50.53 |
| Skin | 37.21 | 31.90 | 34.35 | 100.00 | 40.00 | 57.14 | 86.51 | 33.12 | 47.90 | 100.00 | 38.60 | 55.70 |
| Bladder | 41.67 | 59.59 | 49.04 | 57.14 | 80.00 | 66.67 | 16.11 | 58.11 | 25.23 | 30.19 | 84.21 | 44.44 |
| Heart | 77.21 | 80.93 | 79.03 | 83.33 | 100.00 | 90.91 | 80.73 | 76.33 | 78.47 | 91.00 | 89.22 | 90.10 |
| Adrenal | 20.12 | 7.50 | 10.93 | 100.00 | 33.33 | 50.00 | 2.36 | 10.33 | 3.85 | 13.33 | 66.67 | 22.22 |
| Colon | 23.72 | 13.45 | 17.17 | 33.33 | 33.33 | 33.33 | 1.88 | 14.00 | 3.32 | 20.00 | 33.33 | 25.00 |
| Lymph Node | 56.76 | 39.24 | 46.40 | 100.00 | 40.00 | 57.14 | 5.37 | 22.44 | 8.66 | 7.14 | 11.11 | 8.70 |
| Thyroid | 42.87 | 35.35 | 38.75 | 40.00 | 66.67 | 50.00 | 1.95 | 30.67 | 3.67 | 4.55 | 66.67 | 8.51 |
| Breast | 70.58 | 92.93 | 80.22 | 80.00 | 100.00 | 88.89 | 14.28 | 93.75 | 24.79 | 44.44 | 100.00 | 61.54 |
| Overall | 60.28 | 60.20 | 58.74 | 77.37 | 69.12 | 67.28 | 73.66 | 48.05 | 53.41 | 83.30 | 66.79 | 71.49 |

**Table S2.** Precision, Recall, F1-Score for each organ class separately for test and expanded test set for both patch and slide-level classification using **ResNet50**

| Organ | Test | | | | | | Expanded Test | | | | | |
|---|---|---|---|---|---|---|---|---|---|---|---|---|
| | patch-level | | | slide-level | | | patch-level | | | slide-level | | |
| | Precision | Recall | F1-Score | Precision | Recall | F1-Score | Precision | Recall | F1-Score | Precision | Recall | F1-Score |
| Lung | 15.96 | 20.96 | 18.12 | 100.00 | 20.00 | 33.33 | 89.30 | 31.63 | 46.72 | 95.31 | 48.93 | 64.66 |
| Liver | 83.46 | 48.95 | 61.71 | 75.00 | 60.00 | 66.67 | 73.13 | 55.57 | 63.15 | 89.06 | 72.15 | 79.72 |
| Kidney | 66.62 | 58.85 | 62.49 | 80.00 | 80.00 | 80.00 | 43.58 | 54.33 | 48.37 | 53.04 | 74.39 | 61.93 |
| Brain | 76.98 | 97.81 | 86.15 | 83.33 | 100.00 | 90.91 | 75.19 | 91.14 | 82.40 | 90.32 | 96.55 | 93.33 |
| Spleen | 65.08 | 82.69 | 72.84 | 62.50 | 100.00 | 76.92 | 13.63 | 71.50 | 22.90 | 20.37 | 78.57 | 32.35 |
| Pancreas | 44.18 | 62.90 | 51.90 | 57.14 | 80.00 | 66.67 | 14.78 | 50.11 | 22.83 | 20.34 | 66.67 | 31.17 |
| Prostrate | 49.75 | 75.56 | 60.00 | 38.46 | 100.00 | 55.56 | 32.07 | 62.02 | 42.28 | 39.81 | 91.49 | 55.48 |
| Skin | 48.16 | 28.13 | 35.51 | 100.00 | 60.00 | 75.00 | 91.24 | 34.44 | 50.00 | 100.00 | 52.63 | 68.97 |
| Bladder | 51.87 | 49.83 | 50.83 | 100.00 | 80.00 | 88.89 | 20.97 | 46.95 | 28.99 | 59.09 | 68.42 | 63.41 |
| Heart | 75.15 | 76.69 | 75.91 | 83.33 | 100.00 | 90.91 | 83.43 | 75.33 | 79.18 | 88.35 | 89.22 | 88.78 |
| Adrenal | 16.49 | 7.64 | 10.44 | 0.00 | 0.00 | 0.00 | 1.73 | 10.67 | 2.97 | 0.00 | 0.00 | 0.00 |
| Colon | 26.10 | 12.29 | 16.71 | 50.00 | 33.33 | 40.00 | 1.92 | 11.67 | 3.29 | 20.00 | 33.33 | 25.00 |
| Lymph Node | 43.09 | 26.04 | 32.46 | 100.00 | 20.00 | 33.33 | 3.95 | 14.00 | 6.16 | 7.69 | 11.11 | 9.09 |
| Thyroid | 44.84 | 43.14 | 43.97 | 50.00 | 66.67 | 57.14 | 2.26 | 36.00 | 4.25 | 6.06 | 66.67 | 11.11 |
| Breast | 86.31 | 83.49 | 84.88 | 100.00 | 100.00 | 100.00 | 26.32 | 83.50 | 40.02 | 80.00 | 100.00 | 88.89 |
| Overall | 59.17 | 59.82 | 57.96 | 74.98 | 69.12 | 65.92 | 72.47 | 46.69 | 51.32 | 81.23 | 63.70 | 67.17 |

**Table S3.** Precision, Recall,F1-Score for each organ class separately for test and expanded test set for both patch and slide-level classification using **ViT**

| Organ | Test | | | | | | Expanded Test | | | | | |
|---|---|---|---|---|---|---|---|---|---|---|---|---|
| | patch-level | | | slide-level | | | patch-level | | | slide-level | | |
| | Precision | Recall | F1-Score | Precision | Recall | F1-Score | Precision | Recall | F1-Score | Precision | Recall | F1-Score |
| Lung | 18.62 | 21.46 | 19.94 | 66.67 | 40.00 | 50.00 | 91.56 | 28.60 | 43.59 | 97.59 | 43.32 | 60.00 |
| Liver | 81.88 | 53.56 | 64.76 | 100.00 | 80.00 | 88.89 | 66.62 | 60.62 | 63.48 | 88.41 | 77.22 | 82.43 |
| Kidney | 60.30 | 70.41 | 64.96 | 66.67 | 80.00 | 72.73 | 41.88 | 65.39 | 51.06 | 45.45 | 79.27 | 57.78 |
| Brain | 76.22 | 98.57 | 85.97 | 83.33 | 100.00 | 90.91 | 78.78 | 91.66 | 84.73 | 93.33 | 96.55 | 94.92 |
| Spleen | 62.56 | 72.27 | 67.07 | 57.14 | 80.00 | 66.67 | 11.68 | 62.36 | 19.67 | 18.97 | 78.57 | 30.56 |
| Pancreas | 49.18 | 65.86 | 56.31 | 100.00 | 100.00 | 100.00 | 15.98 | 53.67 | 24.63 | 23.21 | 72.22 | 35.14 |
| Prostrate | 57.65 | 52.08 | 54.72 | 71.43 | 100.00 | 83.33 | 43.55 | 41.81 | 42.66 | 60.00 | 63.83 | 61.86 |
| Skin | 44.63 | 23.83 | 31.07 | 100.00 | 80.00 | 88.89 | 86.73 | 27.15 | 41.35 | 96.43 | 47.37 | 63.53 |
| Bladder | 47.09 | 46.96 | 47.02 | 100.00 | 80.00 | 88.89 | 22.37 | 49.58 | 30.83 | 36.59 | 78.95 | 50.00 |
| Heart | 73.55 | 88.13 | 80.19 | 83.33 | 100.00 | 90.91 | 79.69 | 80.53 | 80.11 | 86.67 | 89.22 | 87.92 |
| Adrenal | 20.70 | 12.54 | 15.62 | 100.00 | 33.33 | 50.00 | 2.10 | 16.67 | 3.72 | 7.69 | 33.33 | 12.50 |
| Colon | 18.99 | 13.30 | 15.65 | 25.00 | 33.33 | 28.57 | 1.75 | 11.00 | 3.02 | 0.00 | 0.00 | 0.00 |
| Lymph Node | 45.19 | 35.43 | 39.72 | 33.33 | 20.00 | 25.00 | 4.60 | 20.33 | 7.50 | 6.45 | 22.22 | 10.00 |
| Thyroid | 46.24 | 48.86 | 47.52 | 75.00 | 100.00 | 85.71 | 2.66 | 46.67 | 5.03 | 8.33 | 100.00 | 15.38 |
| Breast | 88.17 | 85.15 | 86.63 | 100.00 | 100.00 | 100.00 | 29.11 | 82.75 | 43.07 | 66.67 | 100.00 | 80.00 |
| Overall | 58.91 | 60.02 | 58.61 | 78.08 | 76.47 | 75.35 | 72.77 | 46.05 | 49.95 | 81.64 | 60.85 | 64.58 |

**Table S4.** Precision, Recall,F1-Score for each organ class separately for test and expanded test set for both patch and slide-level classification using **Swin Transformer**

| Organ | Test | | | | | | Expanded Test | | | | | |
|---|---|---|---|---|---|---|---|---|---|---|---|---|
| | patch-level | | | slide-level | | | patch-level | | | slide-level | | |
| | Precision | Recall | F1-Score | Precision | Recall | F1-Score | Precision | Recall | F1-Score | Precision | Recall | F1-Score |
| Lung | 21.75 | 26.24 | 23.78 | 100.00 | 40.00 | 57.14 | 89.70 | 35.43 | 50.79 | 97.73 | 57.49 | 72.39 |
| Liver | 83.25 | 46.63 | 59.78 | 100.00 | 60.00 | 75.00 | 68.42 | 57.77 | 62.64 | 88.52 | 68.35 | 77.14 |
| Kidney | 80.88 | 59.94 | 68.85 | 80.00 | 80.00 | 80.00 | 61.71 | 53.40 | 57.26 | 84.42 | 79.27 | 81.76 |
| Brain | 75.35 | 99.56 | 85.78 | 83.33 | 100.00 | 90.91 | 59.25 | 94.62 | 72.87 | 80.00 | 96.55 | 87.50 |
| Spleen | 72.71 | 86.82 | 79.14 | 71.43 | 100.00 | 83.33 | 16.12 | 75.57 | 26.57 | 23.08 | 85.71 | 36.36 |
| Pancreas | 50.27 | 76.79 | 60.77 | 71.43 | 100.00 | 83.33 | 15.97 | 58.67 | 25.11 | 20.63 | 72.22 | 32.10 |
| Prostrate | 47.30 | 65.61 | 54.97 | 44.44 | 80.00 | 57.14 | 31.17 | 57.11 | 40.32 | 32.32 | 68.09 | 43.84 |
| Skin | 51.90 | 22.20 | 31.10 | 100.00 | 40.00 | 57.14 | 92.07 | 20.93 | 34.10 | 100.00 | 22.81 | 37.14 |
| Bladder | 42.64 | 46.83 | 44.64 | 66.67 | 80.00 | 72.73 | 19.28 | 52.37 | 28.19 | 29.55 | 68.42 | 41.27 |
| Heart | 79.17 | 81.96 | 80.54 | 83.33 | 100.00 | 90.91 | 86.12 | 74.36 | 79.81 | 91.00 | 89.22 | 90.10 |
| Adrenal | 22.57 | 13.12 | 16.60 | 100.00 | 66.67 | 80.00 | 2.74 | 20.67 | 4.83 | 14.29 | 33.33 | 20.00 |
| Colon | 25.56 | 12.56 | 16.85 | 50.00 | 33.33 | 40.00 | 1.87 | 10.00 | 3.16 | 0.00 | 0.00 | 0.00 |
| Lymph Node | 52.70 | 38.97 | 44.81 | 100.00 | 40.00 | 57.14 | 5.47 | 22.67 | 8.81 | 8.00 | 22.22 | 11.76 |
| Thyroid | 59.83 | 49.48 | 54.17 | 50.00 | 66.67 | 57.14 | 3.44 | 41.00 | 6.35 | 10.00 | 66.67 | 17.39 |
| Breast | 83.17 | 93.51 | 88.04 | 80.00 | 100.00 | 88.89 | 14.17 | 92.75 | 24.59 | 20.00 | 100.00 | 33.33 |
| Overall | 62.21 | 62.11 | 60.55 | 79.75 | 73.53 | 72.22 | 73.75 | 47.92 | 52.65 | 83.95 | 64.65 | 68.80 |

**Table S5.** Precision, Recall,F1-Score for each organ class separately for test and expanded test set for both patch and slide-level classification using **VGG19**

| Organ | Test | | | | | | Expanded Test | | | | | |
|---|---|---|---|---|---|---|---|---|---|---|---|---|
| | patch-level | | | slide-level | | | patch-level | | | slide-level | | |
| | Precision | Recall | F1-Score | Precision | Recall | F1-Score | Precision | Recall | F1-Score | Precision | Recall | F1-Score |
| Lung | 24.33 | 21.88 | 23.04 | 100.00 | 20.00 | 33.33 | 91.58 | 22.86 | 36.58 | 97.73 | 34.49 | 50.99 |
| Liver | 70.56 | 47.55 | 56.82 | 83.33 | 100.00 | 90.91 | 63.54 | 54.81 | 58.85 | 83.33 | 69.62 | 75.86 |
| Kidney | 61.82 | 63.21 | 62.50 | 80.00 | 80.00 | 80.00 | 43.92 | 61.43 | 51.22 | 53.79 | 86.59 | 66.36 |
| Brain | 70.26 | 98.87 | 82.14 | 71.43 | 100.00 | 83.33 | 51.32 | 94.52 | 66.52 | 67.44 | 100.00 | 80.56 |
| Spleen | 64.14 | 76.36 | 69.72 | 57.14 | 80.00 | 66.67 | 15.30 | 68.71 | 25.03 | 15.63 | 71.43 | 25.64 |
| Pancreas | 48.03 | 42.87 | 45.31 | 100.00 | 80.00 | 88.89 | 17.00 | 31.56 | 22.09 | 39.29 | 61.11 | 47.83 |
| Prostrate | 42.55 | 84.44 | 56.58 | 29.41 | 100.00 | 45.45 | 24.05 | 74.62 | 36.38 | 24.21 | 97.87 | 38.82 |
| Skin | 54.05 | 2.60 | 4.97 | 0.00 | 0.00 | 0.00 | 94.78 | 8.87 | 16.22 | 100.00 | 7.02 | 13.11 |
| Bladder | 65.48 | 33.75 | 44.54 | 100.00 | 60.00 | 75.00 | 22.39 | 29.42 | 25.43 | 66.67 | 31.58 | 42.86 |
| Heart | 67.89 | 80.67 | 73.73 | 62.50 | 100.00 | 76.92 | 70.16 | 76.20 | 73.06 | 78.63 | 90.20 | 84.02 |
| Adrenal | 13.10 | 7.57 | 9.59 | 0.00 | 0.00 | 0.00 | 1.15 | 11.33 | 2.08 | 11.11 | 33.33 | 16.67 |
| Colon | 14.20 | 6.26 | 8.69 | 0.00 | 0.00 | 0.00 | 0.39 | 3.00 | 0.69 | 0.00 | 0.00 | 0.00 |
| Lymph Node | 58.23 | 24.20 | 34.19 | 100.00 | 20.00 | 33.33 | 5.00 | 13.89 | 7.35 | 12.50 | 11.11 | 11.76 |
| Thyroid | 61.93 | 34.56 | 44.36 | 50.00 | 66.67 | 57.14 | 2.47 | 28.00 | 4.54 | 7.69 | 66.67 | 13.79 |
| Breast | 87.13 | 89.25 | 88.18 | 100.00 | 100.00 | 100.00 | 23.88 | 89.25 | 37.68 | 80.00 | 100.00 | 88.89 |
| Overall | 57.28 | 57.91 | 54.97 | 65.72 | 63.24 | 57.95 | 70.05 | 41.83 | 42.84 | 79.53 | 54.69 | 55.24 |

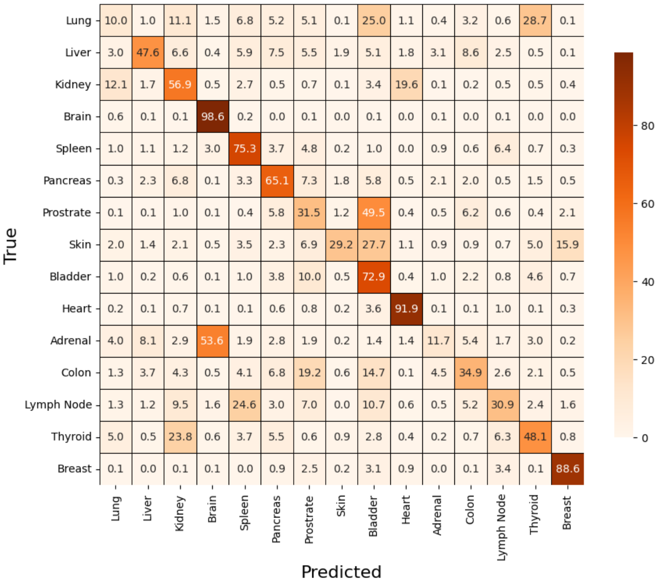


**Figure S17.** Confusion matrix for patch-level classification for test set using **EfficientNet**

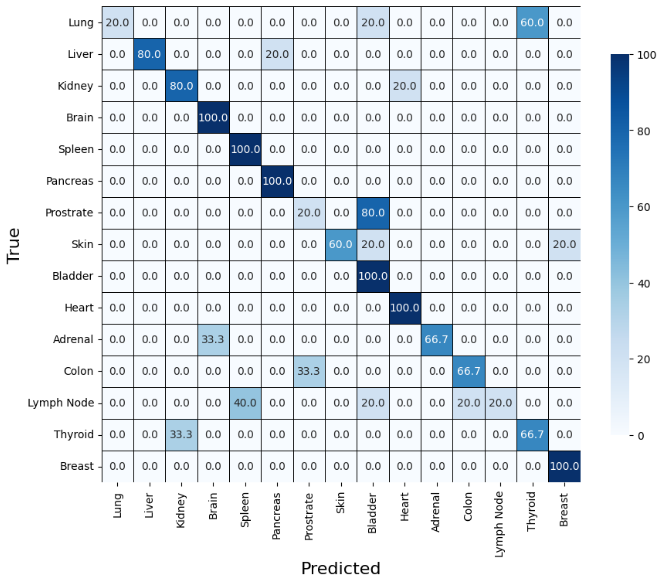


**Figure S18.** Confusion matrix for slide-level classification for test set using **EfficientNet**

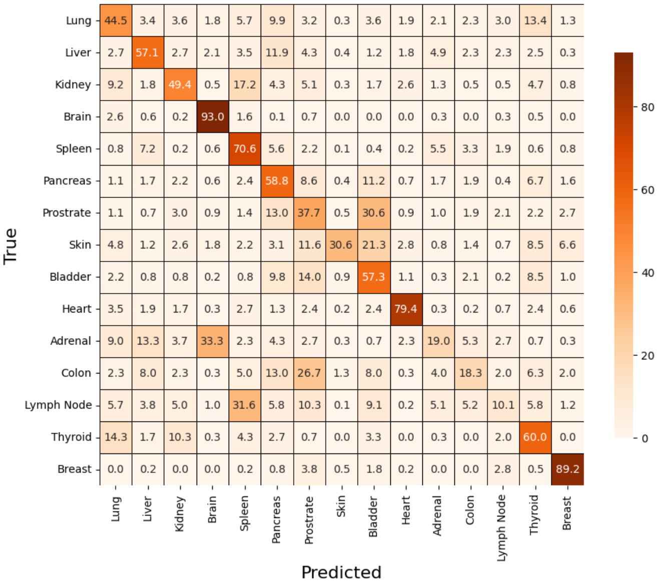


**Figure S19.** Confusion matrix for patch-level classification for expanded test set using **EfficientNet**

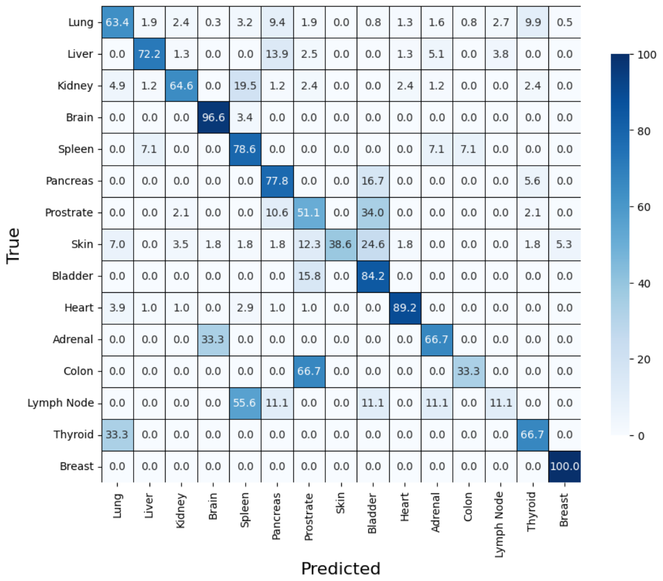


**Figure S20.** Confusion matrix for slide-level classification for expanded test set using **EfficientNet**

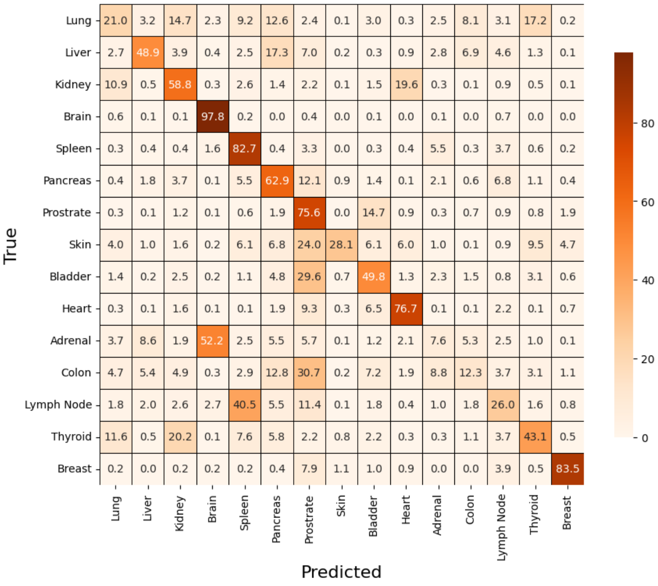


**Figure S21.** Confusion matrix for patch-level classification for test set using **ResNet50**

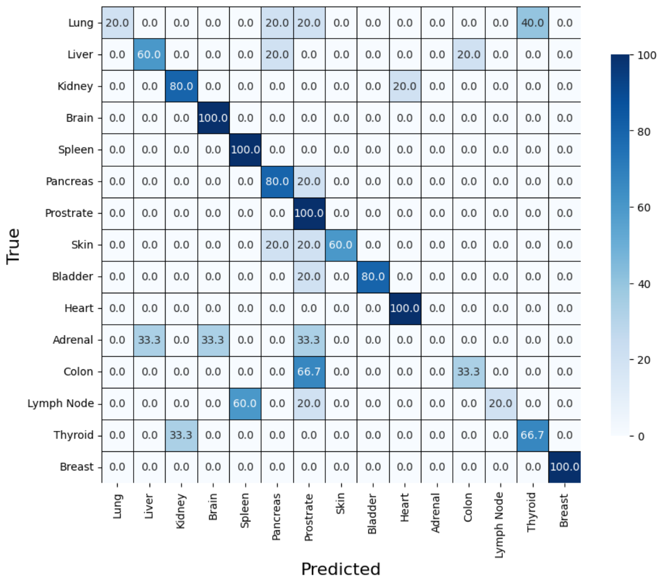


**Figure S22.** Confusion matrix for slide-level classification for test set using **ResNet50**

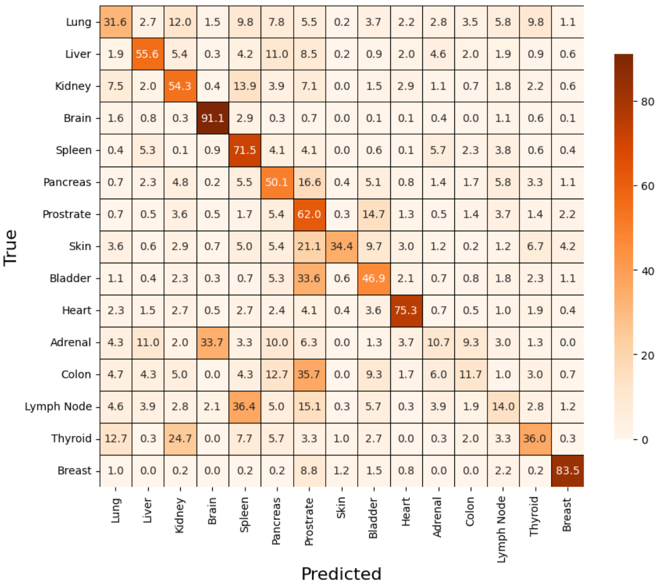


**Figure S23.** Confusion matrix for patch-level classification for expanded test set using **ResNet50**

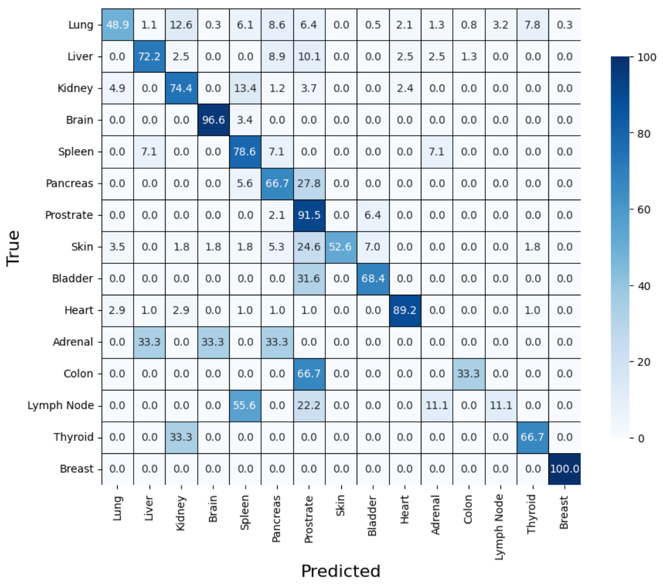


**Figure S24.** Confusion matrix for slide-level classification for expanded test set using **ResNet50**

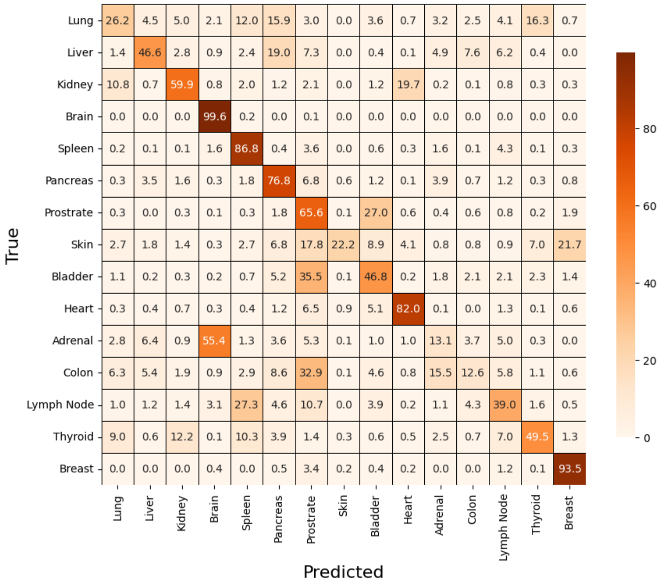


**Figure S25.** Confusion matrix for patch-level classification for test set using **Swin Transformer**

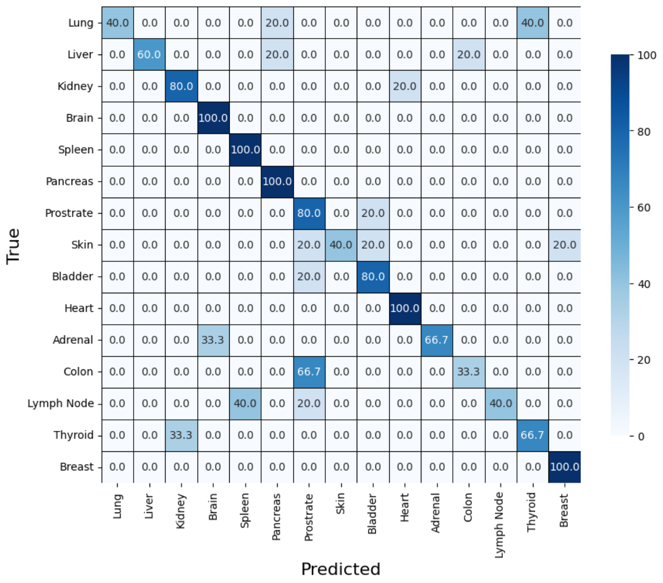


**Figure S26.** Confusion matrix for slide-level classification for test set using **Swin Transformer**

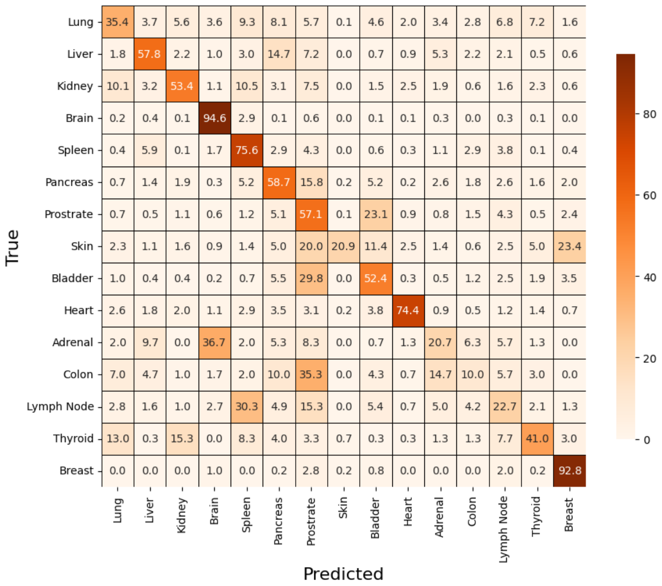


**Figure S27.** Confusion matrix for patch-level classification for expanded test set using **Swin Transformer**

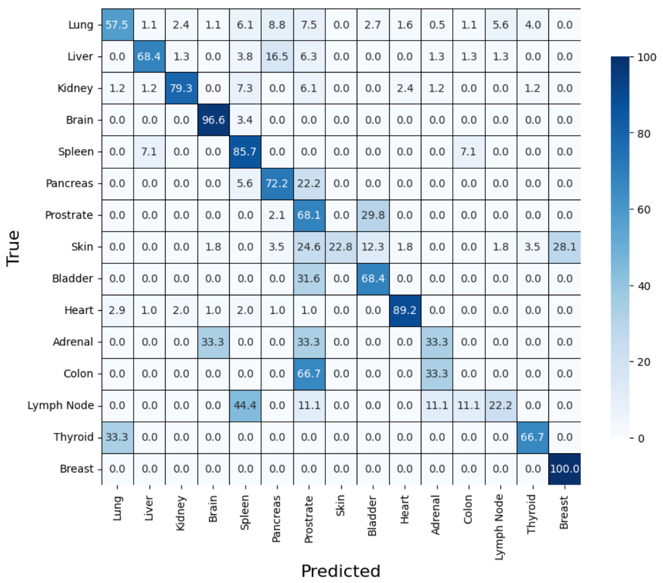


**Figure S28.** Confusion matrix for slide-level classification for expanded test set using **Swin Transformer**

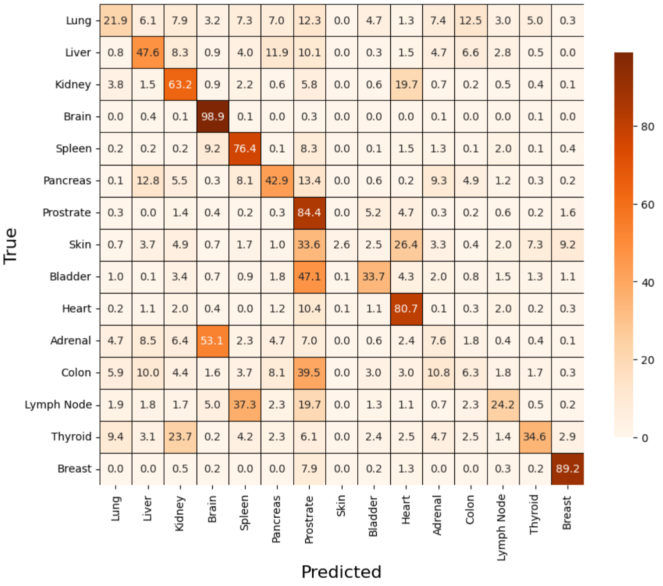


**Figure S29.** Confusion matrix for patch-level classification for test set using **VGG19**

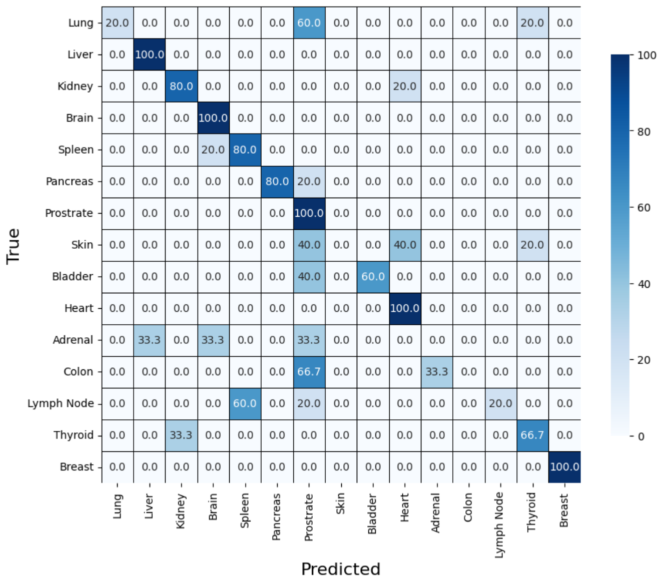


**Figure S30.** Confusion matrix for slide-level classification for test set using **VGG19**

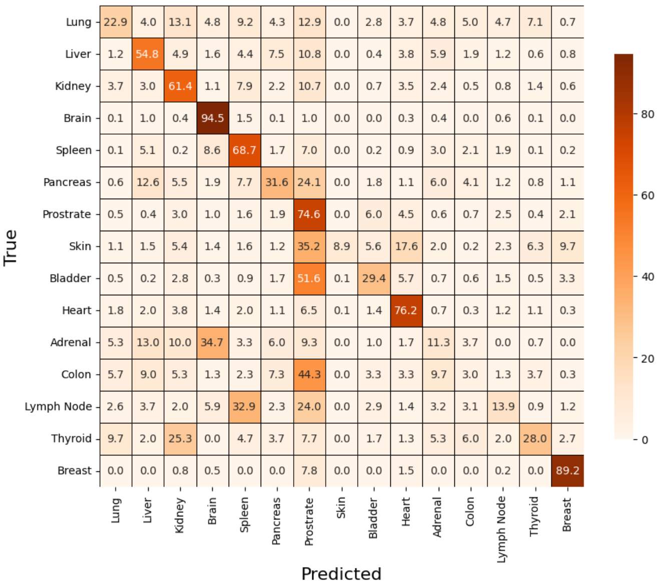


**Figure S31.** Confusion matrix for patch-level classification for expanded test set using **VGG19**

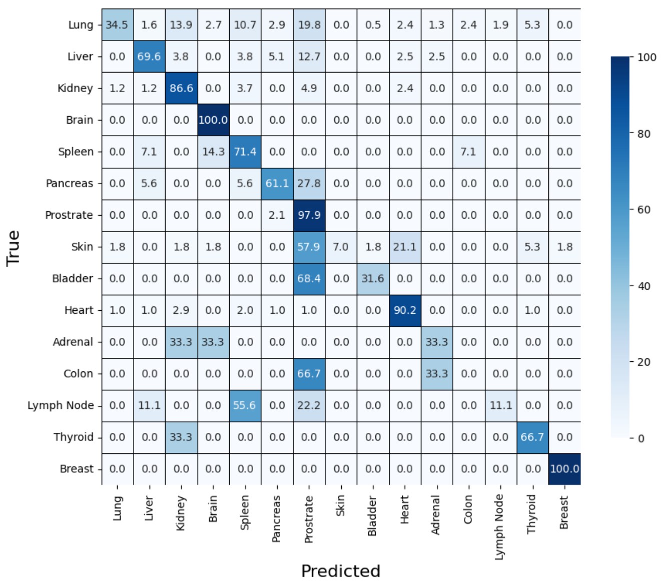


**Figure S32.** Confusion matrix for slide-level classification for expanded test set using **VGG19**
